# Detecting Pathogen-Associated RNA via *Piecewise Isothermal Testing* achieving Sample-to-Result Integration

**DOI:** 10.1101/2021.04.06.21254740

**Authors:** Saptarshi Banerjee, Sujay Kumar Biswas, Nandita Kedia, Rakesh Sarkar, Aratrika De, Suvrotoa Mitra, Subhanita Roy, Aditya Bandopadhyay, Indranath Banerjee, Ritobrata Goswami, Shanta Dutta, Mamta Chawla-Sarkar, Suman Chakraborty, Arindam Mondal

**Affiliations:** School of Bioscience, Indian Institute of Technology Kharagpur, India-721302; School of Medical Science and Technology, Indian Institute of Technology Kharagpur, India-721302; Department of Mechanical Engineering, Indian Institute of Technology Kharagpur, India-721302; B.C. Roy Technology Hospital, Indian Institute of Technology Kharagpur, India-721302; ICMR-National Institute of Cholera and Enteric Diseases, India-700010

**Keywords:** SARS-CoV2, Piecewise Isothermal Nucleic Acid Test, Affordable Diagnostics, Portable Device

## Abstract

We report a novel piece-wise isothermal nucleic acid test (*PINAT*) for diagnosing pathogen-associated RNA that embeds an exclusive DNA-mediated specific probing reaction with the backbone of an isothermal reverse-transcription cum amplification protocol as a unified single-step procedure. This single step sample-to-result test method has been seamlessly integrated in an inexpensive, scalable, pre-programmable and portable instrument, resulting in a generic platform technology for detecting nucleic acid from a wide variety of pathogens. The test exhibited high sensitivity and specificity of detection when assessed using 200 double-blind patient samples for detecting SARS-CoV-2 infection conducted by the Indian Council of Medical Research (ICMR), reporting a positive and negative percent agreement of 94.6% and 98% respectively. We also established its efficacy in detecting influenza-A infection, performing the diagnosis at the point of collection with uncompromised detection rigor. The envisaged trade-off between advanced laboratory-based procedures with the elegance of common rapid tests renders the innovation to be ideal for deployment in resource-limited settings towards catering the needs of the underserved.

## 1. Introduction

The recurrence of epidemics and pandemics from emerging pathogens like Influenza, Ebola, Zika, SARS, MERS and the most recent SARS-CoV-2 has continuously exposed global unpreparedness towards protecting human health, social and economic livelihoods. Considering the possibilities of asymptomatic infected individuals and similarities in symptoms associated with infections caused by various pathogens, common clinical-observation based screening strategies are not adequate to prevent the disease spread. Rather, systematic, aggressive yet accurate testing of individuals on a community level, contact tracing and rapid isolation of infected individuals, when synchronized in tandem, may facilitate an effective management of the epidemic/ pandemic threats. This calls for the development of generic, rapid, affordable yet highly accurate diagnostic tests, which could be easily implemented in resource limited settings.

Different variants of molecular diagnostic tests are central to the detection of pathogenic infections. Point-of-care (POC) immunoassays, alternatively known as rapid antigen or antibody tests, are extremely attractive for deployment in resource limited settings. However, these techniques suffer from limited accuracy owing to rapid genetic evolution of the pathogenic strains and limited abundance of the antigens or antibodies in the body fluid sample, specifically at the early stage of infection. In sharp contrast, the nucleic acid amplification-based tests (NAATs) are intrinsically adapted to capture the most fundamental genomic signature of the infection, and provide a highly sensitive, accurate and reliable approach for diagnosis. Unfortunately, the resource-intensive nature of the laboratory-based procedures for nucleic acid testing (such as real-time quantitative polymerase chain reaction-RT-PCR), limits its large-scale dissemination at de-centralized locations. Isothermal nucleic acid amplification techniques (INAAT), specifically the reverse transcription-loop-mediated isothermal amplification (RT-LAMP), have off-late emerged as potential scalable alternative to RT-PCR (Barreda-García et al., 2018; Foo et al., 2020; Wang et al., 2017, 2016; Xu et al., 2012). The most attractive proposition of RT-LAMP lies in obviating the need of complex thermal cycling protocols so that PCR machines do not remain compulsory. The RT-LAMP reaction is simple, rapid and extremely targeted to identify multiple target regions on the template RNA sequence of the pathogen for amplification and detection. However, despite great promises, the RT-LAMP procedure remains to be extensively applied on a commercial basis, commonly attributed to the constraints such as carryover contamination (Dao Thi et al., 2020) and indirect mode of detection. RT-LAMP reactions are monitored through change in colour (Zhang et al., 2020), turbidity (Tamura et al., 2017) or fluorescence (Shao et al., 2017), which relies upon the change in the environment of the reaction mixture and hence often produces ambiguous results with compromised specificity and sensitivity (Zhang et al., 2020). To overcome this problem, the classical RT-LAMP methods have been recently augmented with the CRISPR-Cas12/13 based detection technology (Broughton et al., 2020; Li et al., 2019), which enhances the overall accuracy of results through sgRNA mediated specific recognition of the cDNA products. However, despite enhancing the specificity of the detection, the inherent instability, and off target effects of sgRNAs point towards the challenges that the CRISPR-based methods may encounter once brought out of highly controlled laboratory settings.

The most compelling challenge that has largely prohibited the INAATs to be deployed for community-level testing is the lack of a simple, user-friendly low-cost instrument that can seamlessly couple any arbitrary isothermal reaction steps customizable with the test protocol. Advancing substantially from NALF (nucleic acid lateral flow) based test-cassettes commercialized by BioHelix (Kolm et al., 2019) and USTAR (Fang et al., 2009), there has, however, been much recent progress in developing fully automated and integrated bench-top NAAT instrumentation for real-time fluorescence detection in less than 2 hours. However, several challenges remain unaddressed for rendering their suitability for community care, including: the need of high voltage power sources (Ramachandran et al., 2020), separate arrangements for cell lysis, image analysis outside the ambit of device integration (Ganguli et al., 2020; Reboud et al., 2019), need of large reagent volume per test (Ganguli et al., 2020), requirement of sphisticated microfabrication steps with inherent complexity, and prohitive cost of the disposable unit (Lafleur et al., 2016; Roskos et al., 2013; Xu et al., 2020), to name a few. Further, none of these provides a generic universal platform for integrated sample-to-result procedure that can be suitably customized as per specific test protocols implementable with minimal manual steps in a resource-limited setting.

Here, we have presented a series of advancements that overcome the aforementioned limitations in the reported nucleic acid based molecular diagnostic procedures. First, we utilized a CRISPR independent but complementary DNA-probe hybridization-based detection procedure integrated with the INAAT protocol to reconstitute a piece-wise isothermal reaction (*abbreviated as PINAT*) with high sensitivity and specificity of detection. The isothermal amplification reaction and the DNA probe hybridization steps could proceed in a concomitant fashion so that the entire process could be performed in a single step closed chamber environment, thus arresting potential cross contamination. In addition, we developed a fully-integrated low-cost yet generic instrument for performing such tests at point-of-use and presented the proof-of-principle for an early prototype. This device comprises a pre-programmable thermal control unit to perform piecewise isothermal reaction steps integrated with a lateral flow assay (LFA) based detection unit. A custom-made mobile app empowered with image analytics and machine learning capabilities was employed for data interpretation, thereby eliminating inaccuracies due to manual interpretation. Our method demonstrated high efficacy in detecting SARS-CoV-2 infection in a double-blind validation process conducted using 200 patient samples with a positive and negative percent agreement of 94.6% and 98% respectively. To extend the generality, we also demonstrated the detection of influenza-A virus RNA directly from infected cells and from virion particles without any additional RNA purification steps.

## 2. Materials and Methods

### 2.1. Portable device fabrication and operation

The design and drawing of the portable instrument was performed using Autodesk Inventor Professional 2021, USA. The outer cover of the device was fabricated using 1.1 mm thick mild steel sheet. The top cover, paper-strip cartridge and microchamber holding cartridges were developed using additive manufacturing technology with a 3D printer (TECH B V30, India). The reaction microchambers were fabricated using transparent polymers. Slots compatible with the sizes of the microchamber units were drilled in the thermal reaction block (heating unit) made of aluminium. All the components required for the development of the thermal control unit were made available from local electronic supplies. The Arduino program was generically developed in-house. The detection unit in the device included a cartridge, holding lateral flow assay (LFA) strips for the visualization via smartphone camera and subsequent analytics. The LFA strips were kept in closed cassettes made of transparent polymer sheets and fabricated using a table-top CNC micro milling machine (T-Tech Inc. USA). (for details see supplementary information).

### 2.2. Test protocol for viral RNA detection

For detecting viral RNA directly from virus particles and infected cells, the samples were heated at 95 °C for 5 minutes prior to the test protocol. The backbone RT-LAMP assay of the integrated protocol was performed using WarmStart® LAMP Kit (DNA & RNA) from New England Biolabs (Cat#M1700L). Six LAMP primers (FIP, BIP, LF, LB, F3, B3) with specific DNA probes (5’ FAM modified) complementary to the loop regions of the LAMP product, named as FLP and BLP, for each gene, were designed and standardized. The overall standardization procedure included heating at 65°C for 30 minutes, 95°C for 5 minutes to inactivate the reaction and then cooling down to 4°C. After addition of the specific DNA probes, the reactions were again heated at 95°C for 5 minutes followed by probe annealing at 50°C for 5 minutes. In single-step sample-to-result integration protocol, the DNA probes (either 3’ modified or 3’ unmodified) were added at the very beginning of the RT-LAMP reaction, with subsequent heating at 65 °C for 30 minutes, 95 °C for 5 minutes to inactivate the reaction and further heating at 50 °C for 5 minutes for probe annealing, seamlessly occurring one after the other without intermediate manual intervention.

### 2.3. DNA based probe hybridization and lateral flow assay

10 picomoles of probe were used per reaction for visualizing the hybridized products on LFA strip. The initial addition of the DNA probe in the RT-LAMP reaction as an integrated single-step protocol, despite being highly favourable for implementation in a point-of-care format with minimal intermediate manual intervention, nevertheless necessitated additional safeguards as this could potentially lead to the concomitant amplification of the probe itself and hence might interfere with the amplification or specificity of the detection process. To avoid such adverse artefacts, we included a double modified DNA probe harbouring 6-FAM and a di-deoxy nucleotide at its 5’- and 3’-termini respectively. Piecewise isothermal reactions were performed in presence of either regular (5’-6-FAM) or dual modified (5’-6-FAM + 3’-ddNTP) DNA probes and subsequently monitored. As evidenced from substantiating experiments, addition of either the 6-FAM or the 6-FMA-3’ddNTP probes from the very beginning of the reaction showed signal comparable to the conventional two-step detection procedure with no non-specific signals in negative control sets. LFA was performed on paper strips from Milenia Biotec (Germany) following the manufacturers protocol. High specificity and inherent stability of the complementary DNA probe ensured high accuracy of the test results even outside the ambit of controlled laboratory ambience.

### 2.4. In-vitro sensitivity assay

Sensitivity assay was performed using 10^4^ to 1 copy of *in vitro* transcribed RNA fragments corresponding to the target gene 1-3. These contrived RNAs were diluted in nuclease free water containing 100ng/ μL or carrier RNA solution for homogeneous distribution of the RNA throughout the solution and hence improved performance of the assay. 1 µl of the diluted RNA was mixed into the reaction master mix and thermal protocol was performed. After implementing the thermal protocol, each reaction was run in 1.5% agarose gel and analysed using lateral flow assay strip. All the reactions were performed in sextuplets. Before performing this sensitivity assay, extensive quality checking of the master mix was performed as mentioned earlier.

### 2.5. Validation using patient samples against gold-standard RT-PCR test

Virus Research and Diagnostic Lab (VRDL), ICMR-NICED, the regional COVID-testing lab in the state of West Bengal, India, receives nasopharyngeal swab samples, suspended in viral transport medium (VTM), of COVID suspected patients referred from hospitals. The clinical samples were tested by Real Time PCR using standard approved kits (ICMR-National Institute of Virology (ICMR-NIV), 2019a, 2019b). RNA was isolated from 200 µl clinical sample using QIAamp Viral RNA kit (Qiagen, Germany). A sample was considered confirmed positive for SARS-CoV-2 infection with a Ct value cut off ≤35. RNase P gene has been used as internal control for all the patient samples. Considering Real Time PCR as Gold Standard, RNA panel of 115 positive samples [equal proportion of high viral load (Ct< 20), moderate (Ct 20-28) and High viral load (Ct 28-35)]; 75 SARS-CoV-2 negative samples (Ct undetermined) and 10 SARS-Cov-2 negative but Influenza A positive (Ct value 20-25) was prepared and coded. The coded panel was transferred to the designated kit validation unit in double-blinded form for validation of the PINAT assay. After the test was run, the panel was de-coded and results of Real Time PCR (Ct values) were compared with PINAT assay.

## 3. Results

### 3.1. A generic portable device for detection of viral infection using INAAT based protocols

The complete implementation workflow from sample collection to result dissemination using the portable device unit is represented in Fig. 1. (1) The swab/ saliva sample collected from the patient is subjected to a brief heating step (described in details in the later sections) performed in heating slots present in the device. (2) The heat lysed sample is subsequently mixed with the test reagents containing primers, probes and enzymes and dispensed altogether in dedicated reaction-chambers (microchambers). (3) The piecewise isothermal time-stamped reactions are held in the portable device without intermediate manual intervention (4) The final products are then dispensed onto the LFA paper strips and the colorimetric readouts developed therein are captured using a smartphone camera, for subsequent image analytics and algorithmic implementation and rapid dissemination of the test outcome.

**Fig. 1.**
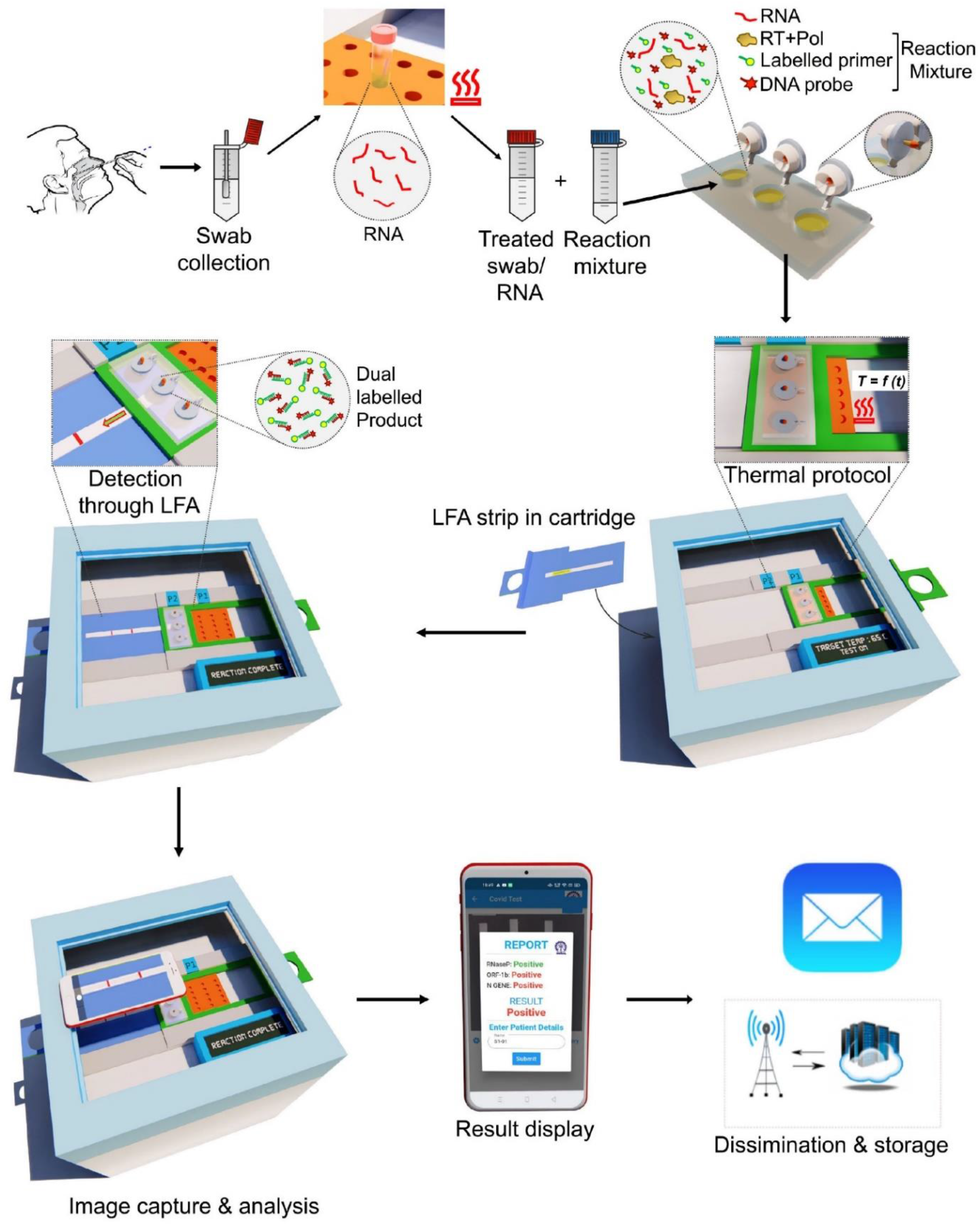
An illustrative overview of PINAT testing for pathogen detection in the integrated device. Followed by collection, the patient swab sample is subjected to a brief initial heat treatment which then can be used for subsequent detection protocol.

The generic point-of-care device developed by us comprises a pre-programmable thermal control cum reaction unit that is capable of performing all the isothermal steps necessary for the overall test protocol. Notably, the same instrument could be customized to execute any arbitrary sequences of isothermal protocols standardized for a given test. The microcontroller based thermal unit of the device includes a heating cartridge, temperature sensor, fan with heat sink assembly, opto-coupler relay (DC) unit that may either trigger heating through cartridge heater or cooling through fan–heat sink assembly and a power unit as switchable alternative. The microcontroller is adapted for selectively carrying out isothermal heating steps including (a) raising temperature to a desired level (b) holding temperature for a specified time and (c) lowering temperature to a desired level. The modular reaction microchambers filled with the test reagents may be placed seamlessly on the heating unit for the intended biochemical reactions. The heating control circuitry and power supply are operatively connected to the heating block with a number of slots that can house the reaction chambers. The numbers of slots can be arbitrarily upscaled to massively parallelize the test procedure.

Once initiated, the entire thermal protocol could be conducted in the closed reaction chambers without any intermediate user intervention, thereby largely arresting the chance of carryover contamination. As a further generic protection against contamination, a needle valve fitted with the reaction chamber was actuated manually to puncture the same at the end of the reaction and dispense the final product seamlessly to the sample pad of a ‘protected’ LFA strip internally encapsulated in the device unit via a movable cartridge. LFA buffer was further dispensed on the strip via a simple dropper to promote capillary-driven transport. The test lines and control lines in the LFA strip were imaged via smartphone camera, analysed and interpreted via mobile app. Smartphone integration eliminated a large number of false negative scenarios by utilizing a dynamic training data set from the previously analysed colorimetric images. The app also facilitates cloud-based data storage and dissemination. Starting from sample collection, the entire test for SARS-CoV-2 detection could be performed within 50 minutes.

The heat lysed sample is mixed with a reaction master mix and dispensed to modular reaction-chambers made of soft polymer tubings for subsequent isothermal amplification process. The right inset shows the caps of the reaction-chamber for air-tight sealing. Piecewise isothermal heating of the reaction-mix occurs in the portable device unit as a single-step process without any intermediate manual intervention. Dispensing of the amplified and labelled cDNA product from reaction chamber to the LFA strip occurs via a manually operated needle valve. Colorimetric detection of the labelled cDNA is made on the LFA strip. A smartphone app based image capture and analysis finally display the result onto the mobile screen that could be disseminated in a controlled fashion through cloud integration.

### 3.2. Standardization of the PINAT protocol for the detection of SARS-CoV-2 genomic RNA

The particular PINAT protocol introduced here for the detection of viral nucleic acid is a generalization and subsequent advancement of the method originally described by Rigano et.al (2014). The method consists of three distinct and extremely generic steps as described in Fig. 2A. First, RT-LAMP reaction in presence of biotinylated forward inner primer (FIP-5’Bt) results in generation of 5’ biotinylated RT-LAMP products. Second, a 6-fluorescein amidite (6-FAM) labelled DNA oligonucleotide (probe), complementary to the loop regions of the RT-LAMP products, is hybridized through consecutive heat denaturation and annealing process thereby generating dual labelled (Biotin + FAM) products. Third, the dual labelled products and the single labelled free probes get separated on a lateral flow assay strip and captured by the streptavidin and secondary anti-FAM antibody immobilized on the test line and control lines, respectively.

**Fig. 2.**
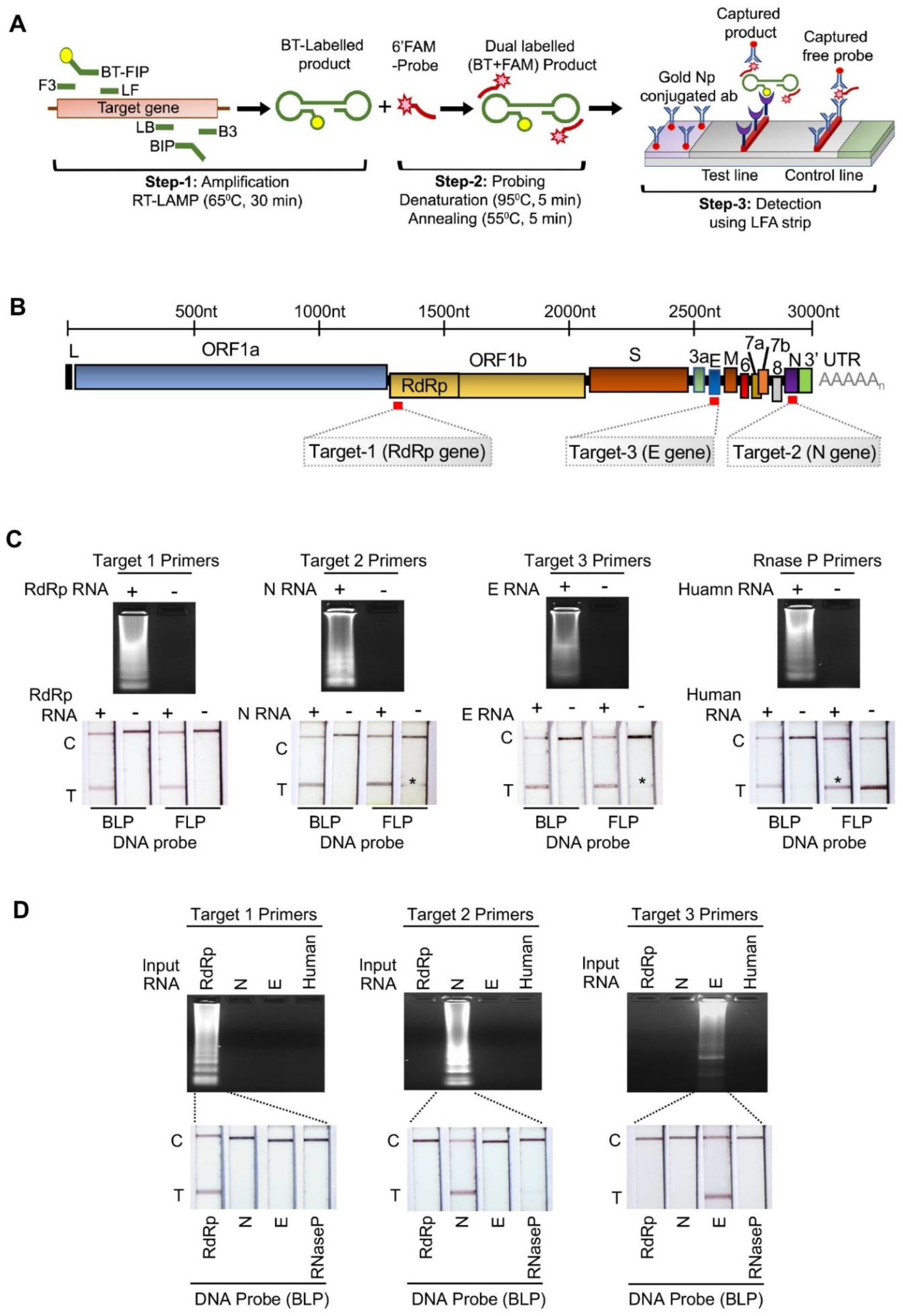
Optimization of the present PINAT test for detecting different target genes in SARS-CoV-2 genomic RNA. (A) Steps of the PINAT based detection of nucleic acid. The target nucleotide sequence was amplified with RT-LAMP primers including biotinylated FIP. The labelled RT-LAMP products were subjected hybridization with FAM labelled complementary DNA probes and detected upon the lateral flow strip. (B) Schematic representation of SARS-CoV-2 genome architecture. Specific target regions were annotated with red bars (C) PINAT-integrated reaction cum detection procedure. Individual target RNAs were amplified through RT-LAMP reaction and analyzed through 2% agarose gel (upper panel). The amplified reactions were subjected to probing with 6-FMA labelled DNA probes (FLP and BLS) and analyzed through paper strip (lower panel). Non-specific appearance of test line in negative control reactions are denoted with asterisks. (D) Cross reactivity of one DNA probe (BLP) towards other SARS-CoV-2 genes or human RNaseP gene.

RT-LAMP primer sets were designed against three highly conserved target regions in the SARS-CoV-2 genomic RNA (Supplementary Fig. S1 & Supplementary Table S2) that reside within the RNA dependent RNA polymerase (RdRp) gene, the nucleocapsid (N) gene and the envelope (E) gene (Fig. 2B). Short in vitro-transcribed (IVT) RNA fragments were used for the standardization of the RT-LAMP reactions (Fig. 2C upper panel). Human RNaseP gene were used as internal control. For complementary DNA probe-based detection via LFA, we designed 6-FAM labelled forward and backward loop probes (FLP and BLP) for each targets and tested specificity. For all target genes, BLPs showed specificity in detection, while, FLPs weak to moderate nonspecific signals (Fig. 2C, lower panel), which could be attributed to partial complementarity of 6-FAM labelled probes with biotin labelled primers, as also supported by our in silico dimer formation analysis. For subsequent experiments, we therefore used BLPs for specific probing and LFA based detection procedure.

RT-LAMP primers designed for one target gene of SARS-CoV-2 showed no cross reactivity against other gene targets (Fig. 2D, upper panel). Additionally, the products amplified from one gene target could only be detected with the corresponding BLPs (Fig. 2D, lower panel), ensuring the high specificity of our double layered detection method. The method also showed no cross reactivity against the genomic RNAs of other corona viruses including SARS, OC43, NL63, 229E and HKU1 (Supplementary Fig. S2B) as well as other RNA viruses like Influenza A, Influenza B or Japanese Encephalitis Virus (JEV) genomic RNAs (Supplementary Fig. S2A) and hence confirmed that it can specifically detect and distinguish SARS-CoV-2 infection from infections caused by other viruses with similar clinical presentations.

In vitro sensitivity assay was performed to ascertain the limit of detection (LOD) of our method for individual target genes. Individual reaction sets containing 10^4^ to 1 copy per microliter of the contrived RNA samples were then subjected to the PINAT reaction in sextuplets. Primers and probes corresponding to all of the gene targets showed the ability to consistently detect 100 copies of RNA molecules in six out of six replicates (Fig. 3A and Supplementary Fig. S3). For the replicates containing 10 copies of RNA, N gene, RdRp and E gene primer sets showed 5/6, 4/6 and 3/6 positive detections, respectively, suggesting the following order of sensitivity: Target 1(RdRP)> Target 2(N)> Target 3(E). All of the gene targets exhibited inconsistent results for higher dilutions suggesting a limit of detection as 10 copies per microliter. Summarily, the PINAT method demonstrated the capability of detecting 10 copies of viral RNA reliably for majority of replicates and 100 copies of viral RNA for all of the replicates, which is comparable to or superior than other RT-LAMP based detection technologies reported in recent times(Broughton et al., 2020; Dao Thi et al., 2020).

**Fig. 3.**
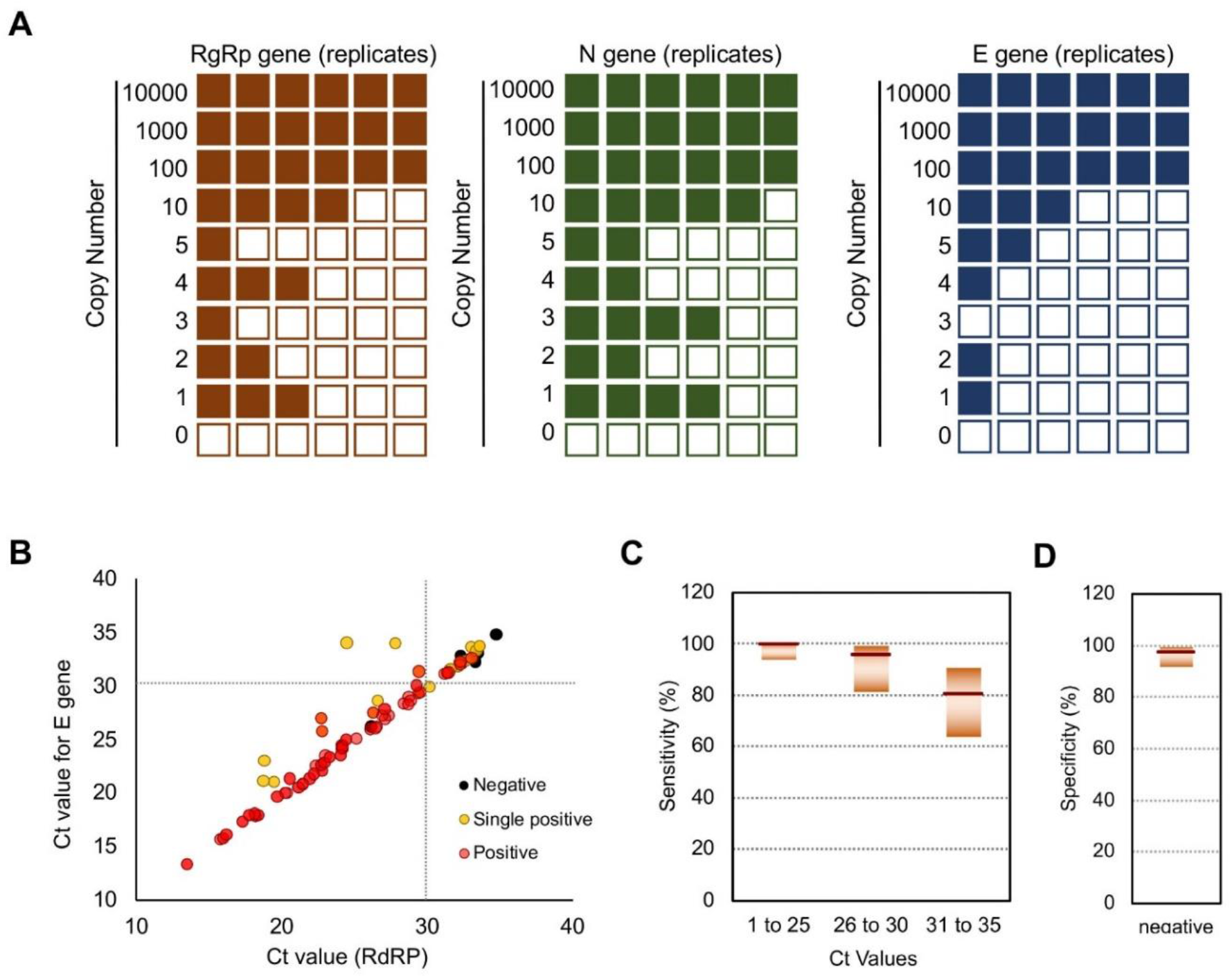
The present PINAT method shows high sensitivity and specificity for detecting SARS-CoV-2 infection in patient samples. (A) Limit of detection of the method for individual gene targets. 10-fold serial dilutions of in vitro synthesized gene fragments were subjected to detection in sextuplets. Coloured boxes represent positive detection while empty boxes represent negative results for individual replicates. (B & C) Patient sample validation results. (B) distribution of the individual samples across the range of Ct values from 25-35. Samples showing positive or negative for both of the gene targets are represented with red and black circles respectively while positive for one and negative for the other gene target are represented with yellow circles. (C) Box plots showing sensitivity of the detection method for patient samples grouped based upon the Ct values. (D) Specificity of the method.

### 3.3. PINAT test for the detection of SARS-CoV-2 infection in patients under resource limited settings

The high specificity and sensitivity of the PINAT test in the in-vitro experiments prompted us to evaluate its efficacy in detecting the presence of SARS-CoV-2 genomic RNA in the nasopharyngeal swab samples isolated from patients. For this purpose, we set up an experimental model which could mimic performing the diagnostic procedure in a remote location in resource limited setting. We pre-aliquoted ready-to-use reaction-mixes in the forms of packed test kits that were transported from our lab to the patient sample testing centre over a road-travel duration of several hours and subsequently stored in a normal refrigerator at 4°C overnight before performing the test procedure (Supplementary Fig. S4 A & B). Purposefully, no specific attention was laid to perform the test in controlled ambience.

We used RNA extracted from 200 double-blinded patient samples, among which 115 were positive and 85 were negative for SARS-CoV-2 infection as determined by the ICMR-NIV developed RT-PCR assay, with the Ct values distributed in between the range of 15 and 35. We employed the primers specific to N gene and RdRp to detect the presence of viral RNA as these two target gene sets showed higher sensitivity than E gene target (Fig. 3A). RNaseP was used as internal control. All the test reactions were performed using the portable instrument and the final results were recorded and analyzed with the help of the custom-made app trained with the algorithm mentioned in Table 1. Briefly, a test with both the gene targets showing positive results were considered as infection positive while negative results with both of them confirmed negative infection. For samples (observed mostly with high CT values) with one gene target showing positive and other showing negative results, the test was repeated and repetition of the exact same trend confirmed positive infection (Fig. 3B). Considering all 200 patient samples, our method showed a positive percentage agreement 93.91% and negative percent agreement 97.64% (Fig. 3C, 3D and Table 2). However, for the samples of Ct values 30 and below, which acts as a suitable reference for clinical decision making, the positive percent agreement values reached to 98.79% (Fig. 3C). To overrule the cross reactivity of the assay, SARS-CoV-2 negative panel (n=85) included 10 known influenza A positive nasopharyngeal samples. All the Influenza positive samples showed negative results for the presence of SARS-CoV-2. This data confirms that the test is capable of detecting SARS-CoV-2 infection in patient samples with high, moderate and low viral loads with superior sensitivity and specificity. Additionally, our experimental model confirms that our test method is suitable to be implemented in the remote areas with limited infrastructure and resources.

**Table 1.**
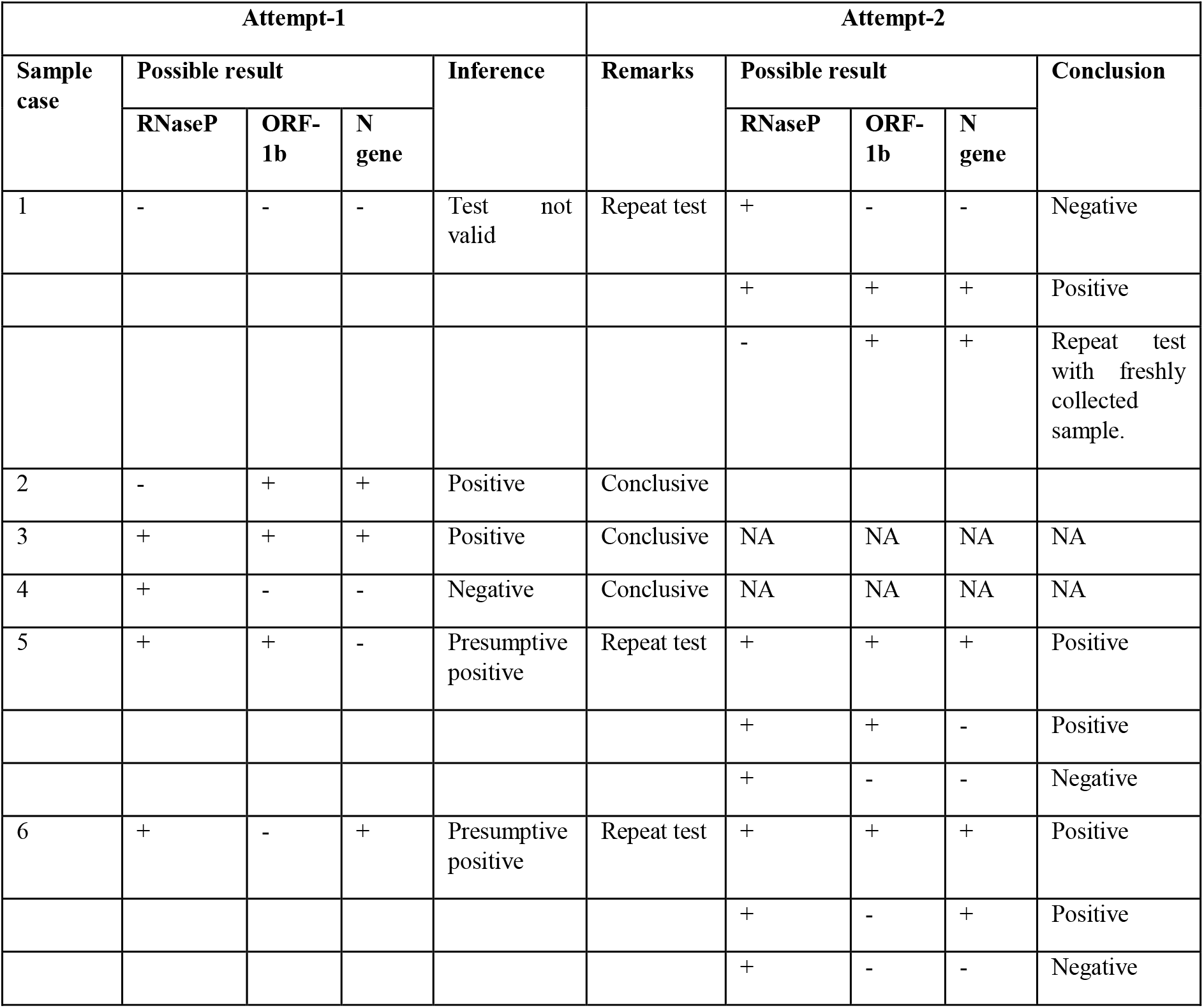
The algorithm for interpretation of the test results.

**Table 2.** PINAT test result for patient-sample based validation of SARS-CoV-2 infection.

|  | Ct value | <= 25 | >25 to 30 | >30 to 35 | Negative |
| --- | --- | --- | --- | --- | --- |
| True | Positive | 59 | 24 | 25 | N/A |
|  | Negative | N/A | N/A | N/A | 83 |
| False | Positive | N/A | N/A | N/A | 2 |
|  | Negative | 0 | 1 | 6 | N/A |
| Sensitivity |  | 100% | 96.15% | 80.65% | N/A |
| Specificity |  | N/A | N/A | N/A | 97.65% |

### 3.4. One step sample-to-solution integration for detection of RNA virus infection

Next, we attempted to standardize a single step swab/ saliva-to-result protocol that could be implemented without changing the basic principle of the PINAT work-flow. For this purpose, we first incorporated an integrated approach for detecting genomic RNA directly from the virus particles or from the infected cells using Influenza A virus as a model system. The PINAT protocol, was standardized for detecting the segments of seven (open reading frame M2) of influenza A/H1N1/WSN/1933 virus genomes (Supplementary Fig. S5). Subsequently, we performed a simulated clinical experiment where influenza A virus infected human lung epithelia cells (A549) were re-suspended in phosphate buffer saline (PBS) spiked with human saliva collected from an infection negative donor. A saliva-PBS suspension containing 100 such cells was then subjected to a brief pre-heating step (95^0^C for 3 minutes) in the portable instrument before being used as an input for the subsequent steps of the PINAT protocol. This adapted protocol was successful in consistently detecting both influenza virus RNA as well as the RNaseP mRNA from the heat ruptured cell lysate either in absence or presence of saliva (0.5% final concentration) (Fig. 4A and Supplementary Fig. S6). Additionally, this protocol was also capable of detecting as low as 500 influenza virion particles (measured in terms of plaque forming units-PFU) diluted in PBS without any additional RNA extraction process in absence of saliva (Fig. 4B and Supplementary Fig. S7). In dilution containing saliva (0.5% final concentration), the method shows a sensitivity of 2500 plaque forming units which is much lower than the usual viral load in a patient showing clinical symptoms(Ward et al., 2004). Similar experiment was performed to ascertain the limit of detection (LOD) for SARS-CoV-2 N gene target in the presence and absence of saliva. Encouragingly, even in presence of saliva, our modified method could successfully detect up to 100 and 10 copies of contrived N gene fragment in 5/6 and 4/6 replicates, which is comparable to the saliva negative controls (Fig. 4C and Supplementary Fig. S8). This ensures high sensitivity and robustness of the N gene target specific primers in amplifying corresponding gene fragment in stringent experimental conditions.

**Fig. 4.**
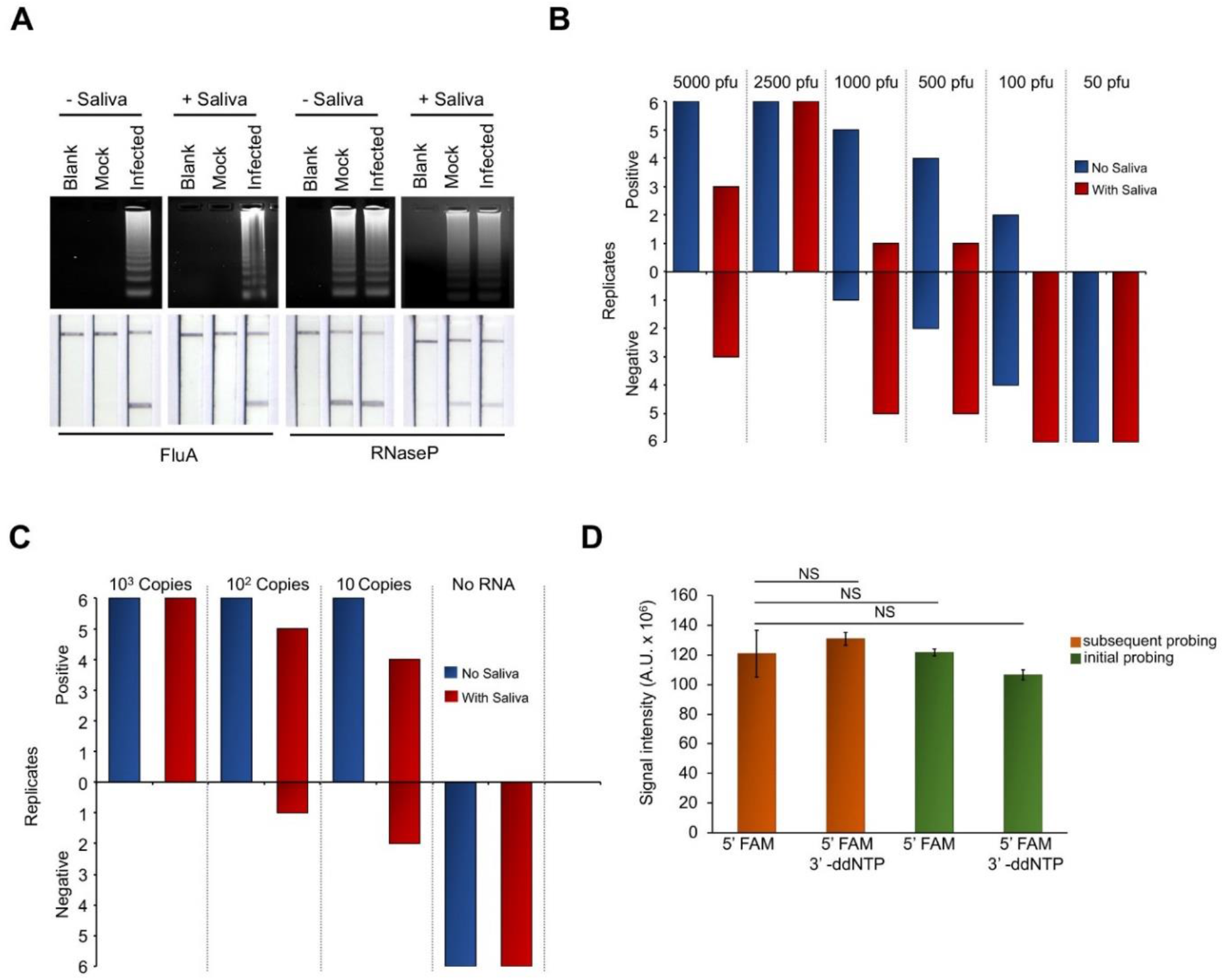
A single step solution for detection of RNA virus infection. (A) Detection of influenza A virus genomic RNA directly from infected cells in the presence or absence of human saliva. Human A549 cells infected with influenza A virus, or mock infected, were resuspended in PBS or PBS spiked with human saliva. The suspension was subjected to the detection procedure without prior RNA extraction procedure, but heating at 95°C for 3 minutes. (B) Defined numbers of influenza virion particles (PFU), amplified from MDCK cells, were subjected to detection procedure (as mentioned in A) in sextuplets either in presence or absence of human saliva. (C) 10-fold serial dilutions of in vitro synthesized N gene fragment were subjected to detection process in sextuplets either in presence or absence of human saliva. (D) Simultaneous amplification and probing for detection of viral RNA in a single step. RdRp Gene fragment was subjected either to RT-LAMP followed by probing in a sequential manner or simultaneous RT-LAMP and probing reaction in a single step process. In both the cases, BLPs labelled with 5’-FAM and with and without 3’-ddNTP modification were used. Products, at the end of the reaction, were subjected to agarose gel electrophoresis in absence of EtBr and imaged under Cy5 channel.

The other exclusive aspect of saliva to result integration, as mentioned earlier, is combining the specific DNA probe hybridization step with the initial amplification step, thereby excluding any in-between manual intervention during the test reactions. For this purpose, we added the 6-FAM labelled complementary DNA probe (BLP) from very beginning of the reaction practically resulting in a master mix which contains all reaction components in it (enzyme mix, RT-LAMP primers, and DNA probe). Hence, mere addition of the input RNA to the master mix and subsequent initiation of the thermal protocol could result in dual labelled products which then could be subjected to detection. As evidenced, addition of either the 6-FAM or the 6-FMA-3’ddNTP probes from the very beginning of the reaction showed signal comparable to the conventional two-step detection procedure with no non-specific signals in negative control sets in either of these cases (Fig. 4D and Supplementary Fig. S9 & S10). Together, this modification along with the saliva-to result integration in the PINAT protocol, when implemented with the help of the instrument innovated by us, holds the potential to revolutionize the nucleic acid-based detection of virus infection and essentially bring high-end molecular diagnostics from sophisticated labs to the field.

## 4. Conclusions and outlook

We developed an integrated highly-accurate one-step nucleic acid test, which is a combination of isothermal amplification, complementary DNA probe hybridization and subsequent detection via lateral flow assay. This method could be performed in a seamless swab-to-result format in a generic inexpensive portable device (∼USD 75.0), thus enabling the scalable deployment of the diagnostics at the point of collection with no discernible compromise in the detection rigor. While relying upon the same basic backbone of isothermal amplification, the present PINAT test attempted to overcome certain practically-inhibiting propositions of the recently-introduced test methods that deploy CRISPR based detection methodology. For instance, the complementary DNA probe-based detection of the present test excludes any reagent-stability concerns as in the case of the sgRNA-mediated detection steps of CRISPR-based technology, when implemented outside controlled laboratory set-up. Furthermore, the DNA probing step can be implemented in a united fashion with the preceding isothermal amplification protocol in contrast to the CRISPR based method where the sgRNA and Cas12/13 complex should be freshly prepared and added after cDNA amplification. This renders the entire PINAT test implementable as a single close tube reaction-sequence without in-between manual intervention, thereby reducing the chance of carryover contamination and the concerned procedural complications to a large extent.

Beyond any specific test protocol, another significant advancement reported here is the generic instrument established as a platform technology which is low cost, user-friendly, customizable and amenable to massive scale-up and parallelization. There is no disposable cartridge except simple paper strips and the reaction chamber, favouring manufacturing scale-up. Each instrument is connectible to secure cloud-based database via smartphone-internet connectivity, enabling a seamless dissemination of the test outcome directly to clinical information systems. The availability of such easy-to-use and reasonably sensitive detection method as a generic platform technology may potentially capture commonly missed instances of early infection and asymptomatic disease presentation and reduce the opportunity for community-level transmission of a large number of diseases. This could result in dramatic improvements in disease management and epidemic/ pandemic control in several unforeseen events in the future. In essence, by exploiting the universal generic structure of innovated test, one may envisage the paradigm of advanced nucleic acid-based diagnostics in the format of a rapid test for several different pathogens. This hybrid paradigm, essentially an amalgamated format with a trade-off between the scientific standards of advanced molecular diagnostics with the elegance of common rapid tests, appears to be the future of diagnostics for the underserved.

## Supporting information

Supplemental protocols, figures, tables

## Data Availability

Detailed data of the patient sample validation work is available in the Supplementary file.

## General

SC and AB acknowledge Sreyansh and Srinivas for helping with the mobile app development.

