## Supplemental protocols, figures, tables for "Detecting Pathogen-Associated RNA via *Piecewise Isothermal Testing* achieving Sample-to-Result Integration"

1. **Details of device fabrication:** The design and drawing of the portable instrument was performed using Autodesk Inventor Professional 2021, USA. The outer cover of the device was fabricated using 1.1 mm thick mild steel sheet to ensure structural robustness without sacrificing economy of manufacturing. The top cover, paper-strip cartridge and microchamber holding cartridges were developed using additive manufacturing technology with a 3D printer (TECH B V30, India). This production is up-scalable via injection moulding for rapid and inexpensive fabrication. The microchamber was fabricated using transparent polymers. Materials like polydimethylsiloxane (PDMA), PMMA and Pyrex were considered for this purpose in different versions of the test instrument, with no perceptible difference in the outcome. The thermal reaction block (heating unit), made of aluminium, was drilled with the numbers of slots same as the numbers of test reactions that may be run in parallel. This aspect of the design is completely flexible and hallmarked by unrestricted scalability, at the expense of increasing the size of the device. In-house and validation experiments were conducted with maximum 30 numbers of such reaction slots, which could be accommodated, along with the LFA strip based detection unit, altogether in a compact cuboidal box with linear dimensions restricted within 1 foot. All the components required for the development of the thermal control unit and digital display were made available from local electronic supplies and were subsequently assembled. The Arduino program was generically developed in-house for performing arbitrary piece-wise isothermal heating steps necessary for any chosen reaction protocol. The thermal unit

was tested extensively to minimize the fluctuation from the set temperature by the controller for a target temperature to be achieved in the heat block. The thermal unit was integrated with the detection unit in the same portable instrument. The detection unit included a cartridge holding LFA strips for the visualization of the test lines and control lines via smartphone camera and subsequent analytics as well as dissemination of the test outcome by the same smartphone. The LFA strips were kept in closed cassettes made of transparent polymer sheets and fabricated using a table-top CNC micro milling machine (T-Tech Inc. USA). For carrying out the test, the cartridge holding the microchamber filled with the reaction-mix and/ or test sample was first placed on the heating slots of the thermal unit. Subsequently, the device was switched on for executing the PINAT steps one after the other as pre-programmed as per the specific protocol to be implemented, without requiring intermediate manual intervention. Upon completion of the reaction, the same cartridge was pushed forward to an identified location in the instrument for seamless dispensing of the products on the LFA strip for the subsequent detection step. A needle valve was actuated for dispensing of the final reaction product straight onto the sample pad of the LFA (see Supplementary video). The test results were visualized and analyzed via smartphone app. The method and implementation was designed and tested to be invariant across different smartphone versions with minor adjustment.

2. **Bioinformatics analysis**: Multiple sequence alignment was performed by using the NCBI virus database align utility. Total 500 complete genome sequence was selected (Supplementary Table S3). Aligned file was viewed in Bio Edit Sequence alignment editor. To analyze the conservation of each position, positional numerical summary was calculated in Bio Edit software. Gap features present only in the reference sequence was omitted in the analysis. Number of recurrences of nucleotide at a specific position, identical to the reference sequence, or the nucleotide present mostly at a specific position was taken into consideration. The specific number was divided by the total number of sequence and converted into percentage. A completely conserved sequence showed 100% value for the calculation. The complete dataset was plotted graphically in Microsoft Excel software. To check the conservation of the specific primers annealing sites, the same multiple alignment file was used. Regions corresponding to primers or their annealing site were selected and submitted to WebLogo website (https://weblogo.berkeley.edu/logo.cgi) for generation of Logo Plot.

3. **Statistical analysis**: Positive percent agreement and negative percent agreement was calculated using the following formulae:

$$\text{Positive Percent Agreement} = \frac{\text{T.P}}{\text{T.P+F.N}} \times 100$$

$$\text{Negative Percent Agreement} = \frac{\text{T.N}}{\text{T.N+F.P}} \times 100$$

Where T.P, T.N, F.P, F.N denote true positive, true negative, false positive and false negative respectively. 95% confidence interval was calculated for the data set using Wilson's method.

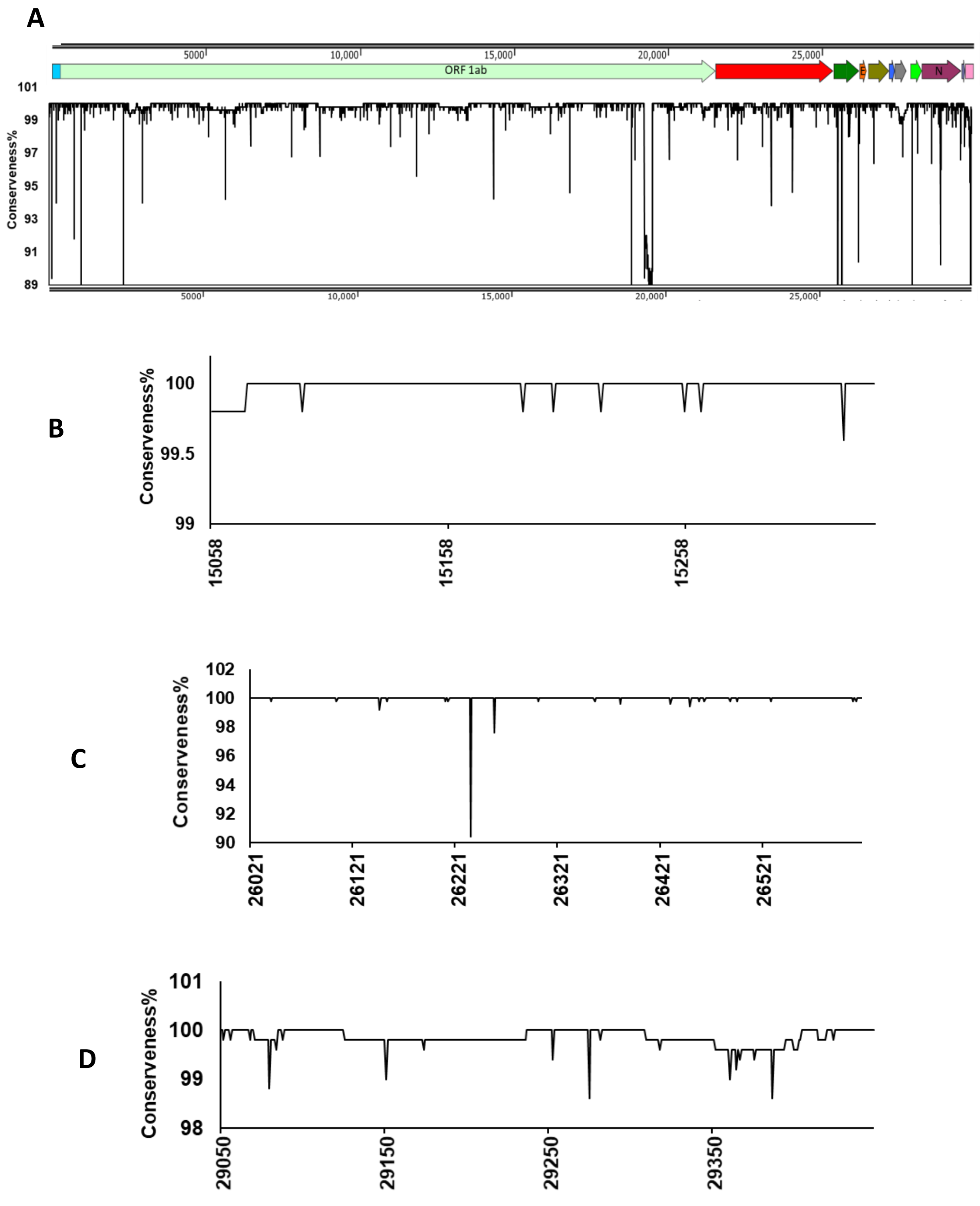


**Supplementary Fig. S1: Analysis of the conserved nature of the SARS CoV-2 genome.** (A) Total 500 complete sequences (listed in supplementary Table S1) were aligned with the reference sequence. Positions with gap in reference sequence were not considered. For each position, conservation was calculated by analyzing the positional numerical summery in Bioedit software. Conserved nature of each target regions are expanded in (B) RdRP, (C) N and (D) E.

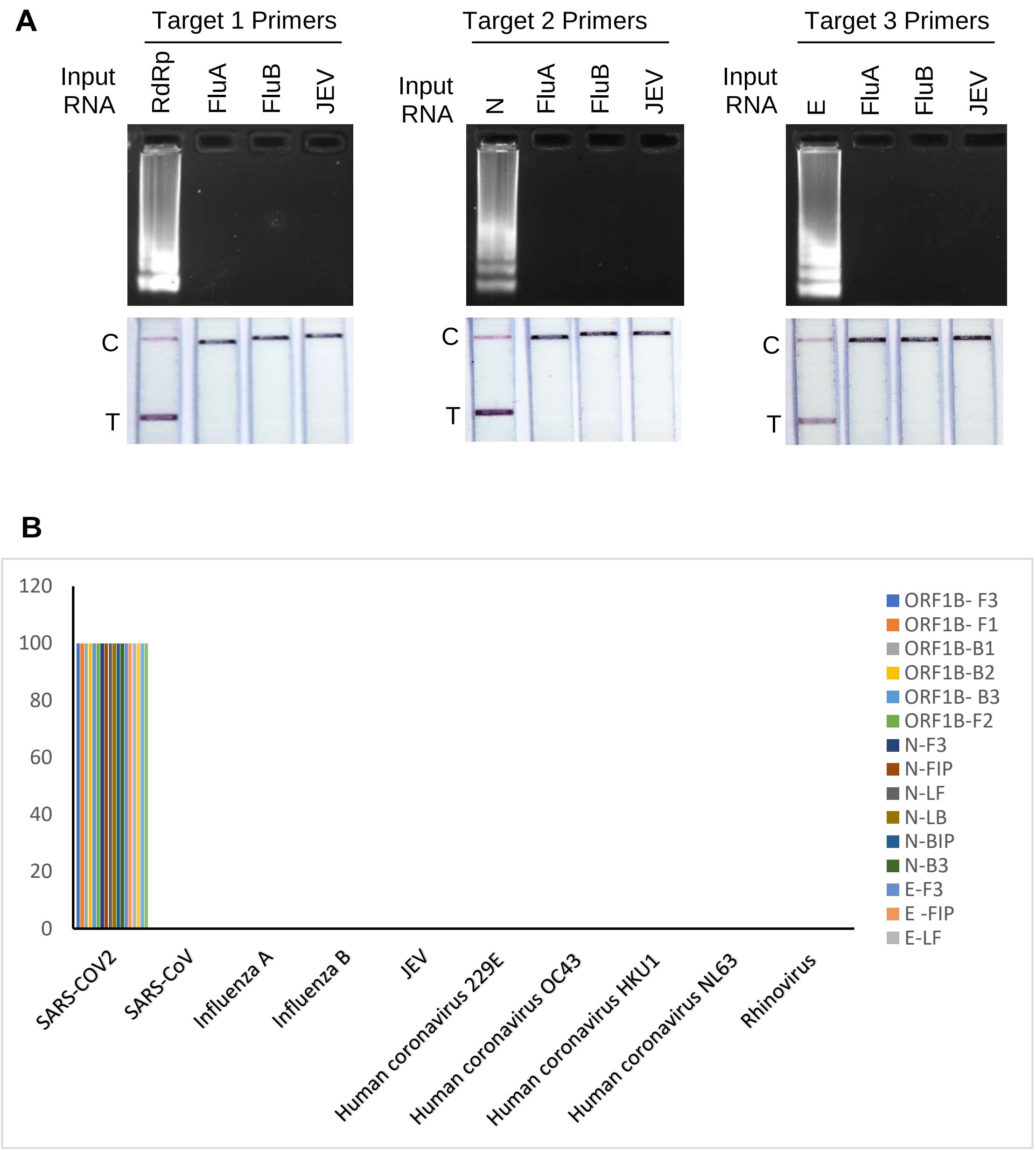


**Supplementary Fig. S2**: **Cross reactivity of SARS-CoV-2 primers against the genomic RNA of other viruses. (A)** Cross reactivity of COVID-19 specific primers were tested using PINAT protocol against influenza A, influenza B and Japanese Encephalitis viral genomic RNAs isolated from virus particles. Reaction products were analyzed through LFA based detection. Respective SARS-CoV-2 target genes were used as positive control. **(B)** Cross reactivity of SARS-CoV-2 specific primers were tested against different RNA viruses in silico.

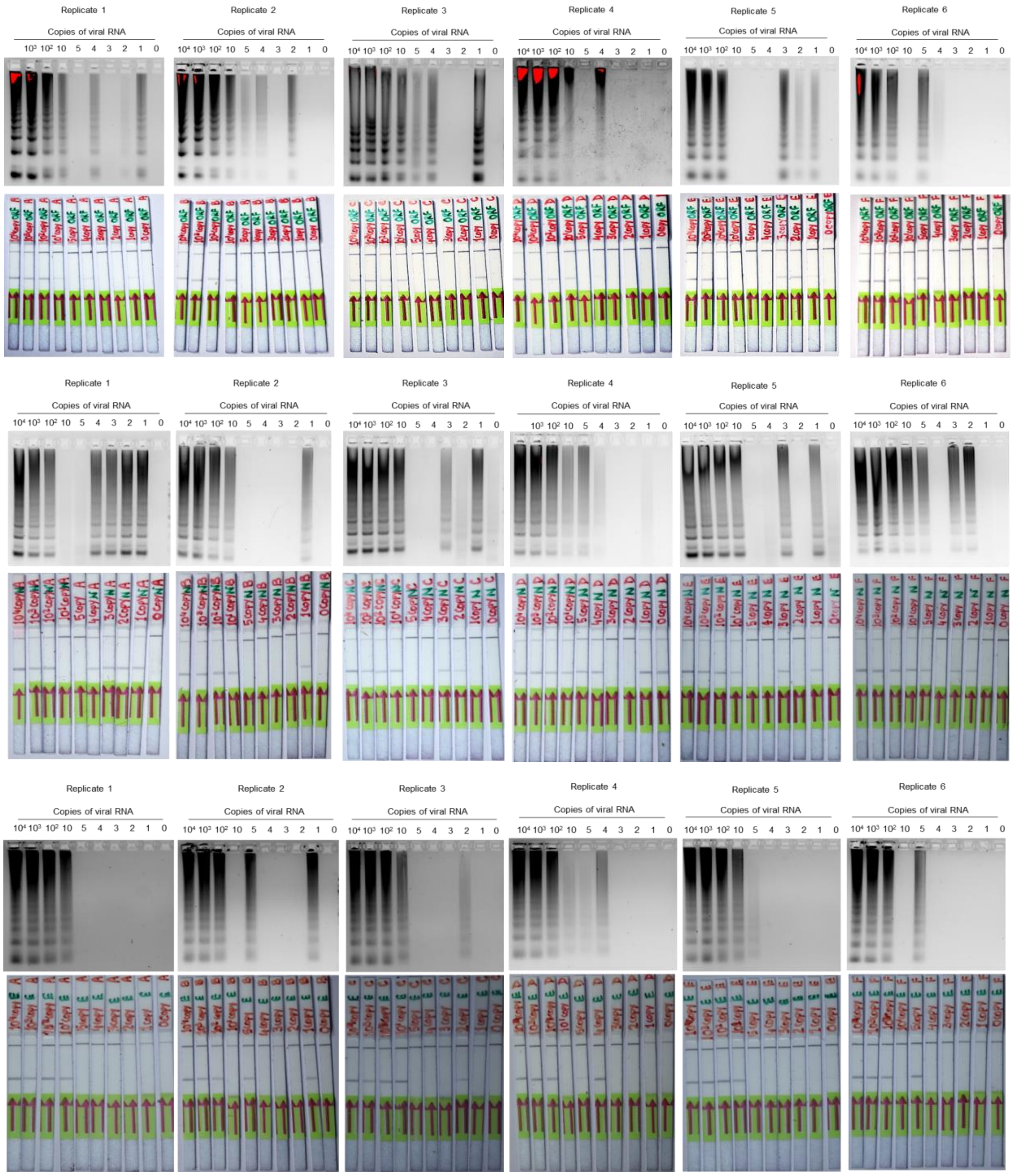


**Supplementary Fig. S3**: **Sensitivity of PINAT using SARS-CoV-2 specific primers and probe (COVIRAP test).** PINAT protocols were performed with ten-fold serial dilutions of the gene fragment in sextuplets. RT-LAMP reaction products were analyzed by agarose gel electrophoresis followed by DNA probe hybridization and LFA based detection (upper panel–RdRP specific primers, middle panel-N gene specific primers and lower panel-E gene specific primers)

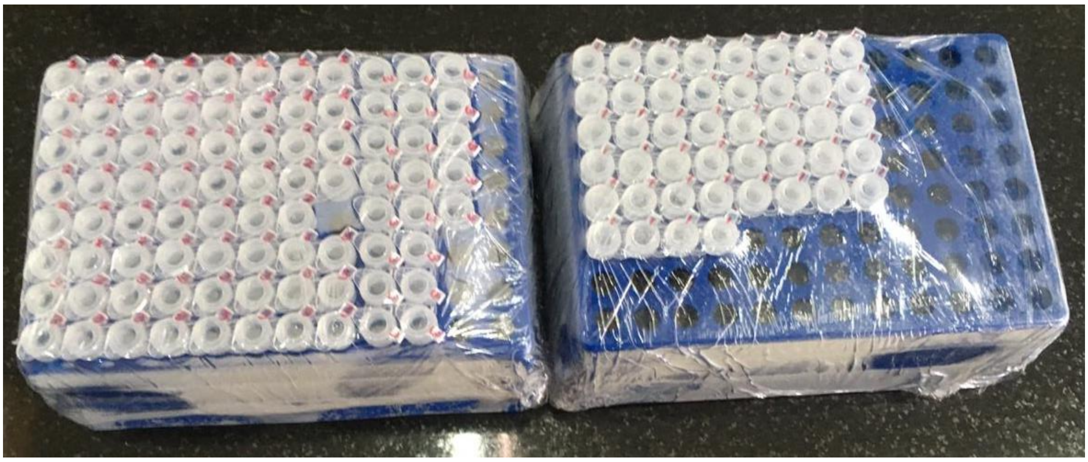

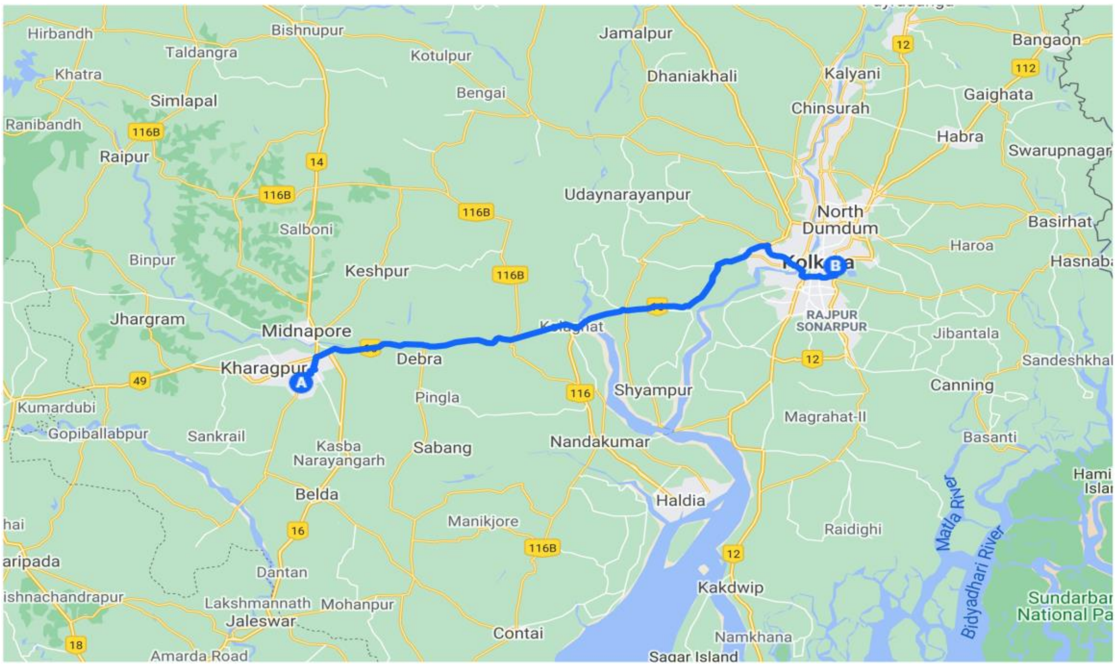


**Supplementary Fig. 4**: Kit packaging (upper) and transportation pathway to the test center (lower). The reaction master mixes corresponding to N and RdRp gene targets were reconstituted, tested for quality control and aliquoted in reaction tubes. The reaction master mixes were then transported in ice packs from Indian Institute of Technology Kharagpur to ICMR-NICED where the test with patient samples were conducted using the test method. In peak traffic hours, the nearly 150 km of road travel used to take around 4.5 hours.

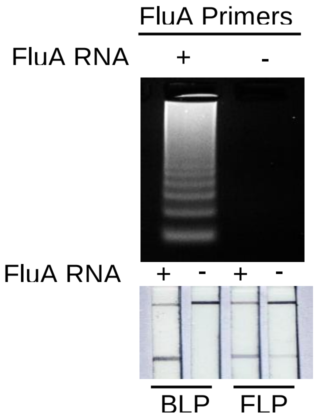


**Supplementary Fig. S5**: PINAT for detection of influenza virus RNA extracted from viruses amplified in MDCK cells. RT-LMAP products were analyzed through agarose gel electrophoresis followed by hybridization with backward or forward loop probed (BLP, FLP) and detected on lateral flow assay strip.

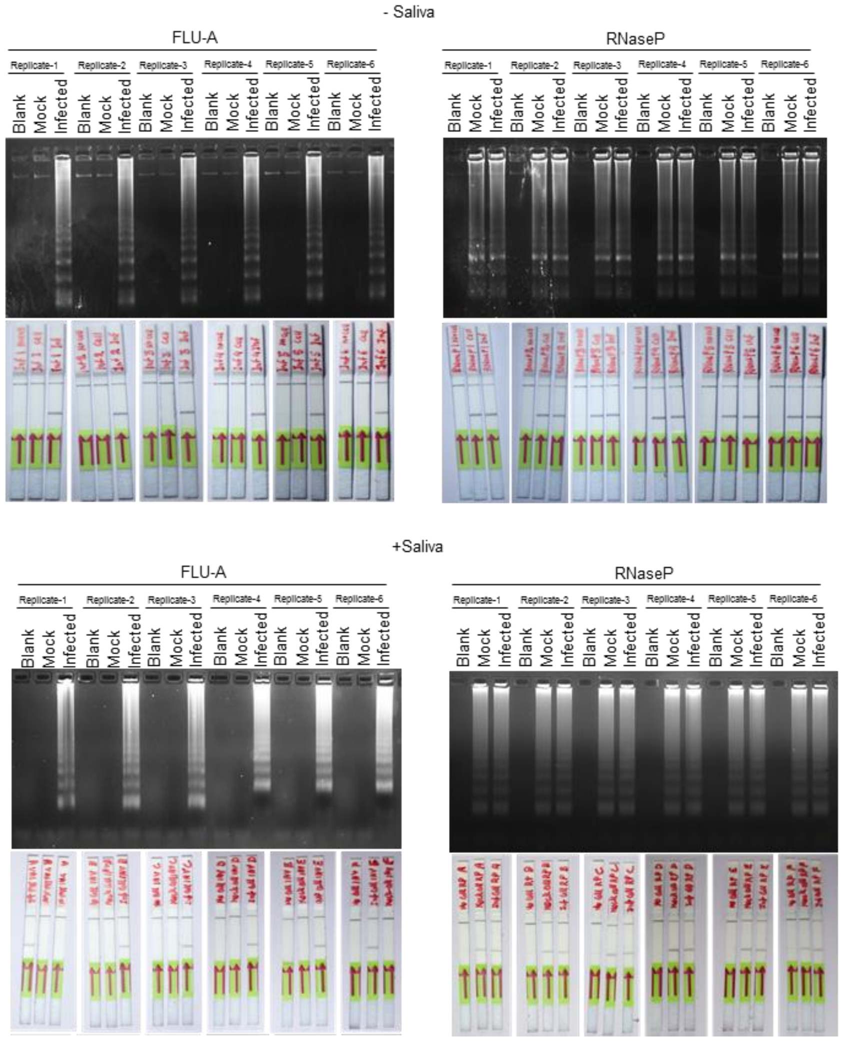


**Supplementary Fig. S6**: PINAT for detection of influenza virus RNA directly from infected A549 cells without any RNA purification. A549 cells, either infected with influenza A virus or mock infected, were resuspended in PBS mixed or PBS spiked with human saliva were subjected to heating at 95$^{0}$C for 3 minutes before using as an input RNA for the PINAAT protocol. RT-LAMP reaction products were analyzed by agarose gel electrophoresis followed by DNA probe hybridization and LFA based detection. Blank sets contains no cells, but only PBS or PBS spiked with human saliva.

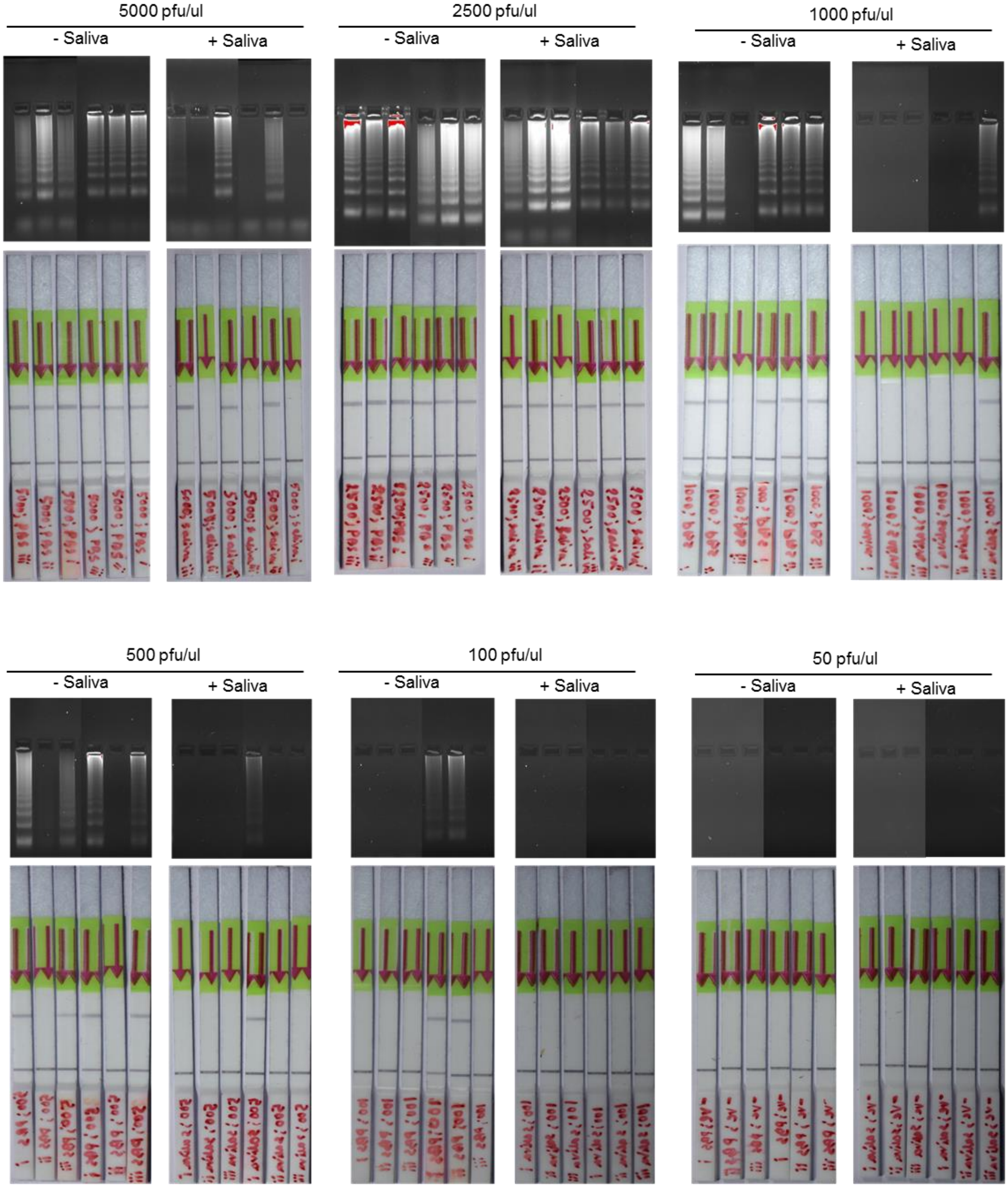


**Supplementary Fig. S7**: Limit of detection of PINAT for detecting influenza A virus RNA directly from virus particles in presence and absence of human saliva. Known PFU of influenza virus particles were diluted in PBS or PBS spiked with human saliva. Subsequently, the sample was directly used for the PINAT test (including prior heating at $95^{0}$C for 3 minutes) without RNA purification.

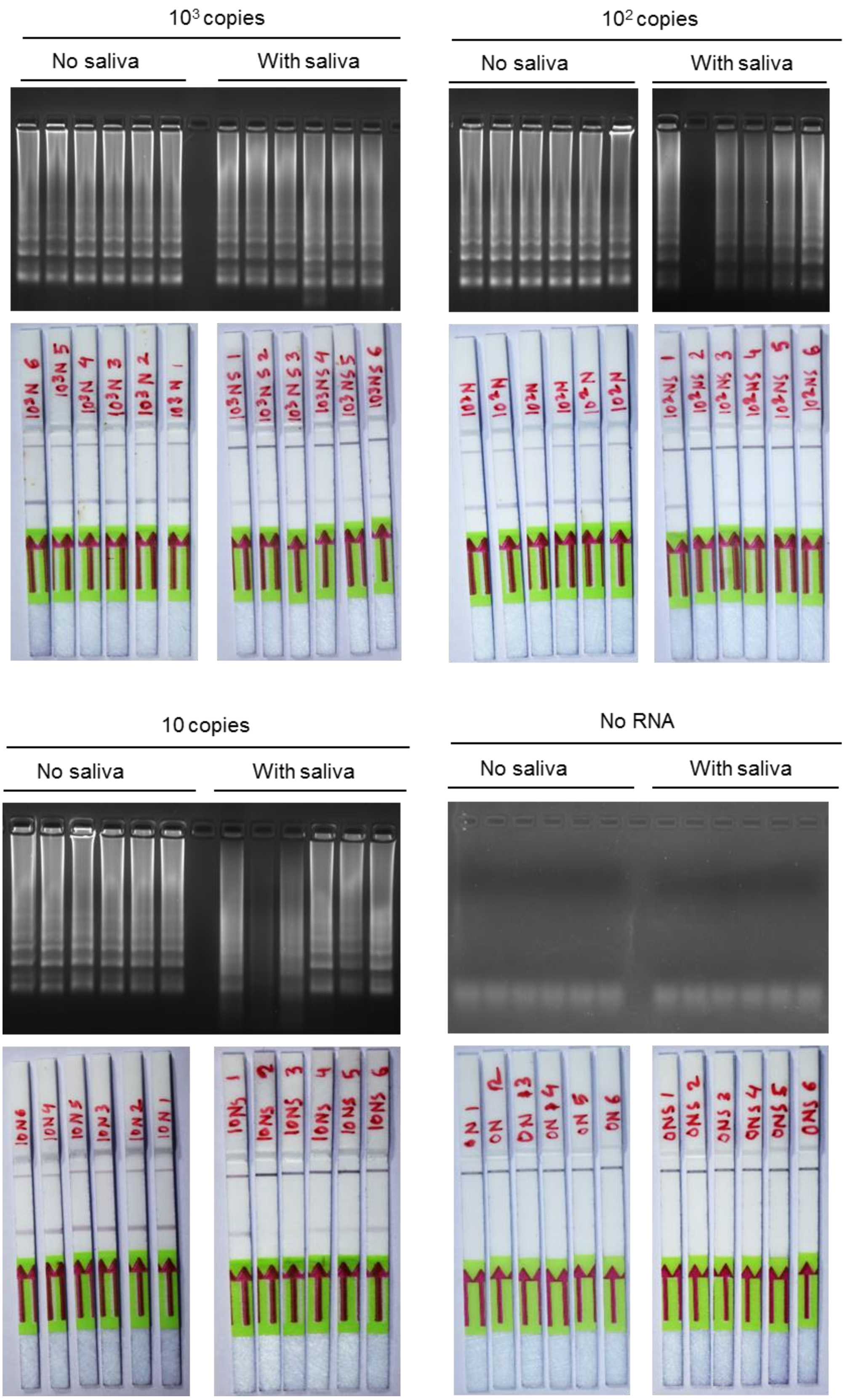


**Supplementary Fig. S8**: Sensitivity of PINAT for detecting SARS-CoV-2 N-gene specific RNA in presence of human saliva sample. 10-fold serial dilutions of the in-vitro transcribed SARS-CoV-2 N-gene RNA were prepared in PBS or PBS spiked with human saliva which were then subjected to the PINAT protocol after heating at 95$^0$C for 3 minutes.

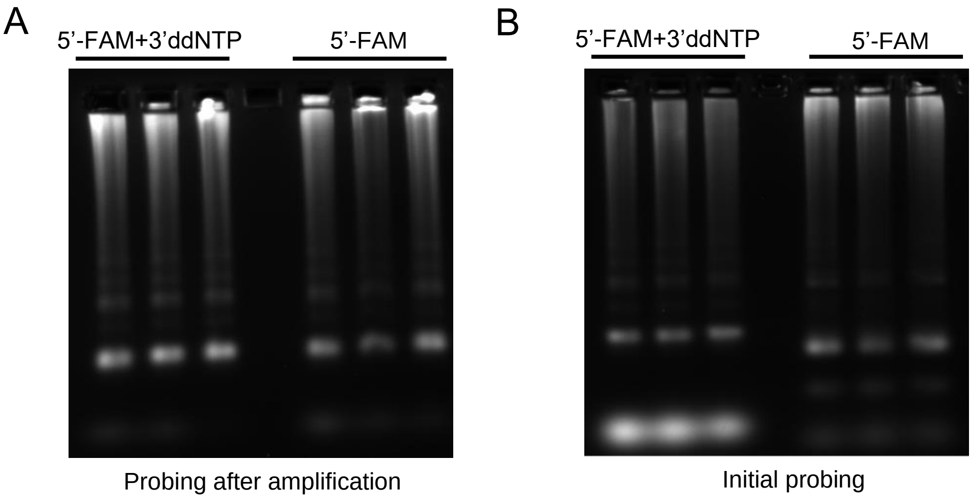


**Supplementary Fig. S9**: (A) RT-LAMP reactions were performed with in vitro transcribed SARS-CoV-2 N-gene target region and the corresponding primers. Products were subjected to hybridization with BLPs labelled with 5’ FAM and with or without 3’ ddNTP modification. (B) For the integrated amplification-cum-probing step-based protocol, both kinds of modified BLPs were added at the beginning of RT-LAMP reaction followed by amplification, heat denaturation and hybridization steps carried out without any interruptions. Products were analyzed through agarose gel electrophoresis in absence of EtBr and imaged under fluorescein channel.

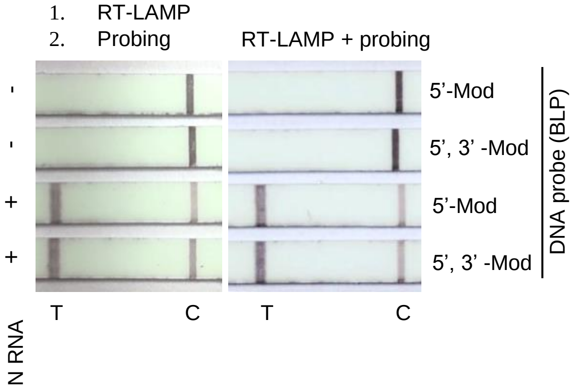


**Supplementary Fig. S10**: RT-LAMP reactions were performed with in vitro transcribed SARS-CoV-2 N-gene target region and the corresponding primers. Products were subjected to hybridization with BLPs labelled with 5’ FAM and with or without 3’ ddNTP modification (left panel). For the integrated amplification-cum-probing step-based protocol, both kinds of modified BLPs were added at the beginning of RT-LAMP reaction followed by amplification, heat denaturation and hybridization steps carried out without any interruptions. Products were analysed through LFA.

**Supplementary Table S1:** Details of patient sample validation results

| | **FAM** | **HEX** | **ROX** | **Result** | **ROUND 1** | | | | **ROUND 2** | | | | **Final Result** | **REMARKS** |
|---|---|---|---|---|---|---|---|---|---|---|---|---|---|---|
| **Sample Number** | **E gene** | **ORF1ab** | **Rnase P** | | **Rnase p** | **RdRP** | **N gene** | **Result** | **Rnase P** | **RdRP** | **N gene** | **Result** | | |
| KP1 | **28.71** | **29.01** | **26.25** | **Positive** | + | + | + | Positive | | | | | **Positive** | Concordant |
| KP2 | **27.34** | **27.27** | **26.77** | **Positive** | + | + | + | Positive | | | | | **Positive** | Concordant |
| KP3 | **26.06** | **26** | **25.86** | **Positive** | + | + | + | Positive | | | | | **Positive** | Concordant |
| KP4 | **26.53** | **26.24** | **24.19** | **Positive** | + | + | + | Positive | | | | | **Positive** | Concordant |
| KP5 | **26.42** | **26.08** | **24.62** | **Positive** | + | + | + | Positive | | | | | **Positive** | Concordant |
| KP6 | N/A | N/A | 28.16 | **Negative** | + | - | - | Negative | | | | | **Negative** | Concordant |
| KP7 | N/A | N/A | 28.11 | **Negative** | + | - | - | Negative | | | | | **Negative** | Concordant |
| KP8 | N/A | N/A | 30.43 | **Negative** | + | - | - | Negative | | | | | **Negative** | Concordant |
| KP9 | N/A | N/A | 30.2 | **Negative** | + | - | - | Negative | | | | | **Negative** | Concordant |
| KP10 | **26.13** | **26.21** | **30.18** | **Positive** | + | - | - | Negative | | | | | Negative | Discordant |
| KP11 | **27.07** | **26.94** | **25.74** | **Positive** | + | + | + | Positive | | | | | **Positive** | Concordant |
| KP12 | **24.41** | **24.99** | **18.57** | **Positive** | + | + | + | Positive | | | | | **Positive** | Concordant |
| KP13 | **32.61** | **32.36** | **23.66** | **Positive** | + | - | + | REPEAT | + | - | + | Positive | **Positive** | Concordant |
| KP14 | N/A | N/A | 27.86 | **Negative** | + | - | - | Negative | | | | | **Negative** | Concordant |
| KP15 | N/A | N/A | 28.35 | **Negative** | + | - | - | Negative | | | | | **Negative** | Concordant |
| KP16 | N/A | N/A | 32.12 | **Negative** | + | - | - | Negative | | | | | **Negative** | Concordant |
| KP17 | N/A | N/A | 28.94 | **Negative** | + | - | - | Negative | | | | | **Negative** | Concordant |
| KP18 | N/A | N/A | 28.17 | **Negative** | + | - | - | Negative | | | | | **Negative** | Concordant |
| KP19 | **31.19** | **31.14** | **26.93** | **Positive** | + | + | + | Positive | | | | | **Positive** | Concordant |
| KP20 | N/A | N/A | 30.18 | **Negative** | + | - | - | Negative | | | | | **Negative** | Concordant |
| KP21 | **32.11** | **31.83** | **27.05** | **Positive** | + | + | - | REPEAT | + | + | - | Positive | **Positive** | Concordant |
| KP22 | N/A | N/A | 28.17 | **Negative** | + | + | - | REPEAT | + | - | - | Negative | **Negative** | Concordant |
| KP23 | N/A | N/A | 28.02 | **Negative** | + | + | + | Positive | | | | | Positive | Discordant |
| KP24 | N/A | N/A | 27.94 | **Negative** | + | - | - | Negative | | | | | **Negative** | Concordant |
| KP25 | **28.35** | **28.4** | **31.24** | **Positive** | + | + | + | Positive | | | | | **Positive** | Concordant |
| KP26 | **28.85** | **28.65** | **30.6** | **Positive** | + | + | + | Positive | | | | | **Positive** | Concordant |
| KP27 | **31.35** | **31.24** | **26.78** | **Positive** | + | + | - | REPEAT | + | + | - | Positive | **Positive** | Concordant |
| KP28 | N/A | N/A | 27.9 | **Negative** | + | - | - | Negative | | | | | **Negative** | Concordant |

| | | | | | | | | | | | | | | |
|---|---|---|---|---|---|---|---|---|---|---|---|---|---|---|
| KP29 | N/A | N/A | 30.99 | **Negative** | + | - | - | Negative | | | | | **Negative** | Concordant |
| KP30 | N/A | N/A | 27.4 | **Negative** | + | - | - | Negative | | | | | **Negative** | Concordant |
| KP31 | N/A | N/A | 32.4 | **Negative** | + | - | - | Negative | | | | | **Negative** | Concordant |
| KP32 | N/A | N/A | 27.35 | **Negative** | + | - | - | Negative | | | | | **Negative** | Concordant |
| KP33 | **29.38** | **29.32** | **32.2** | **Positive** | + | + | + | Positive | | | | | **Positive** | Concordant |
| KP34 | **34.72** | **34.81** | **28.08** | **Positive** | + | - | - | Negative | | | | | Negative | Discordant |
| KP35 | **30.14** | **29.94** | **27.14** | **Positive** | + | - | + | REPEAT | + | - | + | Positive | **Positive** | Concordant |
| KP36 | **32.24** | **32.03** | **29.92** | **Positive** | + | - | + | REPEAT | + | - | + | Positive | **Positive** | Concordant |
| KP37 | N/A | N/A | 29.67 | **Negative** | + | - | - | Negative | | | | | **Negative** | Concordant |
| KP38 | N/A | N/A | 28.06 | **Negative** | + | - | - | Negative | | | | | **Negative** | Concordant |
| KP39 | **33.3** | **32.28** | **30.1** | **Positive** | + | - | - | Negative | | | | | Negative | Discordant |
| KP40 | **32.28** | **32.24** | **29.29** | **Positive** | + | + | - | REPEAT | + | + | - | Positive | **Positive** | Concordant |
| KP41 | **32.27** | **32.82** | **29.75** | **Positive** | + | - | - | Negative | | | | | Negative | Discordant |
| KP42 | **32.39** | **32.19** | **29.57** | **Positive** | + | - | - | Negative | | | | | Negative | Discordant |
| KP43 | **33.01** | **33.65** | **27.61** | **Positive** | + | + | - | REPEAT | + | + | - | Positive | **Positive** | Concordant |
| KP44 | **33.49** | **33.11** | **27.26** | **Positive** | + | - | - | Negative | | | | | Negative | Discordant |
| KP45 | **33.04** | **32.63** | **27.71** | **Positive** | + | + | - | REPEAT | + | + | - | Positive | **Positive** | Concordant |
| KP46 | **33.27** | **32.78** | **27.16** | **Positive** | + | - | - | Negative | | | | | Negative | Discordant |
| KP47 | **33.35** | **33.29** | **27.31** | **Positive** | + | + | - | REPEAT | + | + | - | Positive | **Positive** | Concordant |
| KP48 | **33.6** | **33.74** | **27.3** | **Positive** | + | - | + | REPEAT | + | - | + | Positive | **Positive** | Concordant |
| KP49 | **31.58** | **31.56** | **26.32** | **Positive** | + | + | - | REPEAT | + | + | - | Positive | **Positive** | Concordant |
| KP50 | **31.45** | **31.28** | **25.61** | **Positive** | + | + | + | Positive | | | | | **Positive** | Concordant |
| KP51 | N/A | N/A | 28.21 | **Negative** | + | - | - | Negative | | | | | **Negative** | Concordant |
| KP52 | N/A | N/A | 27.73 | **Negative** | + | - | - | Negative | | | | | **Negative** | Concordant |
| KP53 | N/A | N/A | 29.44 | **Negative** | + | - | + | REPEAT | + | - | - | Negative | **Negative** | Concordant |
| KP54 | N/A | N/A | 27.02 | **Negative** | + | - | - | Negative | | | | | **Negative** | Concordant |
| KP55 | N/A | N/A | 28.4 | **Negative** | + | + | - | REPEAT | + | - | - | Negative | **Negative** | Concordant |
| KP56 | **29.41** | **31.36** | **29.15** | **Positive** | + | + | - | REPEAT | + | + | - | Positive | **Positive** | Concordant |
| KP57 | **26.58** | **28.61** | **22.72** | **Positive** | - | + | + | Positive | | | | | **Positive** | Concordant |
| KP58 | **27.05** | **27.87** | **25.48** | **Positive** | + | + | + | Positive | | | | | **Positive** | Concordant |
| KP59 | **22.69** | **26.97** | **22.8** | **Positive** | + | + | + | Positive | | | | | **Positive** | Concordant |
| KP60 | **27.8** | **34.01** | **29.55** | **Positive** | + | + | - | REPEAT | + | + | - | Positive | **Positive** | Concordant |
| KP61 | N/A | N/A | 28.39 | **Negative** | + | + | - | REPEAT | + | - | - | Negative | **Negative** | Concordant |

| | | | | | | | | | | | | | | |
|---|---|---|---|---|---|---|---|---|---|---|---|---|---|---|
| KP62 | N/A | N/A | 28.18 | Negative | + | + | + | Positive | | | | | Positive | Discordant |
| KP63 | N/A | N/A | 29.49 | Negative | + | + | - | REPEAT | + | - | - | Negative | Negative | Concordant |
| KP64 | N/A | N/A | 30.22 | Negative | + | - | - | Negative | | | | | Negative | Concordant |
| KP65 | N/A | N/A | 27.53 | Negative | + | - | - | Negative | | | | | Negative | Concordant |
| KP66 | N/A | N/A | 28.45 | Negative | + | + | - | REPEAT | + | - | - | Negative | Negative | Concordant |
| KP67 | N/A | N/A | 32.21 | Negative | + | - | - | Negative | | | | | Negative | Concordant |
| KP68 | N/A | N/A | 30.49 | Negative | + | - | + | REPEAT | + | - | - | Negative | Negative | Concordant |
| KP69 | N/A | N/A | 27.29 | Negative | + | - | - | Negative | | | | | Negative | Concordant |
| KP70 | N/A | N/A | 29.93 | Negative | + | - | - | Negative | | | | | Negative | Concordant |
| KP71 | 26.25 | 27.5 | 25.47 | Positive | + | + | - | REPEAT | + | + | - | Positive | Positive | Concordant |
| KP72 | 32.21 | 32.2 | 29.63 | Positive | + | + | - | REPEAT | + | + | - | Positive | Positive | Concordant |
| KP73 | 31.39 | 31.22 | 31.78 | Positive | + | + | - | REPEAT | + | + | - | Positive | Positive | Concordant |
| KP74 | N/A | N/A | 28.94 | Negative | + | - | - | Negative | | | | | Negative | Concordant |
| KP75 | N/A | N/A | 27.78 | Negative | + | - | - | Negative | | | | | Negative | Concordant |
| KP76 | 26.88 | 27.23 | 30.02 | Positive | + | + | + | Positive | | | | | Positive | Concordant |
| KP77 | 28.69 | 28.29 | 28.63 | Positive | + | + | + | Positive | | | | | Positive | Concordant |
| KP78 | N/A | N/A | 27.12 | Negative | + | - | - | Negative | | | | | Negative | Concordant |
| KP79 | N/A | N/A | 27.98 | Negative | + | - | + | REPEAT | + | - | - | Negative | Negative | Concordant |
| KP80 | N/A | N/A | 28.76 | Negative | + | - | - | Negative | | | | | Negative | Concordant |
| KP81 | 38.99 | N/A | 30.27 | Negative | + | - | + | REPEAT | + | - | - | Negative | Negative | Concordant |
| KP82 | 39.75 | N/A | 31 | Negative | + | - | - | Negative | | | | | Negative | Concordant |
| KP83 | 24.14 | 24.48 | 27.7 | Positive | + | + | + | Positive | | | | | Positive | Concordant |
| KP84 | 24.18 | 24.35 | 27.57 | Positive | + | + | + | Positive | | | | | Positive | Concordant |
| KP85 | 39.39 | N/A | 29.29 | Negative | + | + | - | REPEAT | + | - | - | Negative | Negative | Concordant |
| KP86 | 38.66 | N/A | 26.97 | Negative | + | - | - | Negative | | | | | Negative | Concordant |
| KP87 | N/A | N/A | 29.23 | Negative | + | - | - | Negative | | | | | Negative | Concordant |
| KP88 | 24.15 | 24.28 | 27.55 | Positive | + | + | + | Positive | | | | | Positive | Concordant |
| KP89 | 39.38 | N/A | 30.65 | Negative | + | - | + | REPEAT | + | - | - | Negative | Negative | Concordant |
| KP90 | 18.78 | 23.03 | 30.63 | Positive | - | + | + | Positive | | | | | Positive | Concordant |
| KP91 | 22.78 | 25.76 | 26.04 | Positive | + | + | + | Positive | | | | | Positive | Concordant |
| KP92 | 36.27 | N/A | 28.42 | Negative | + | - | - | Negative | | | | | Negative | Concordant |
| KP93 | N/A | N/A | 31.63 | Negative | + | - | - | Negative | | | | | Negative | Concordant |
| KP94 | 24.44 | 34.05 | 25.72 | Positive | + | - | + | REPEAT | + | - | + | Positive | Positive | Concordant |

| | | | | | | | | | | | | | | |
|---|---|---|---|---|---|---|---|---|---|---|---|---|---|---|
| KP95 | **19.45** | **21.04** | **23.58** | **Positive** | + | - | + | REPEAT | + | - | + | Positive | **Positive** | Concordant |
| KP96 | **22.35** | **22.56** | **23.47** | **Positive** | + | + | + | Positive | | | | | **Positive** | Concordant |
| KP97 | N/A | N/A | 23.75 | **Negative** | + | - | - | Negative | | | | | **Negative** | Concordant |
| KP98 | **24.13** | **24.14** | **29.21** | **Positive** | + | + | + | Positive | | | | | **Positive** | Concordant |
| KP99 | **18.71** | **21.12** | **21.71** | **Positive** | + | - | + | REPEAT | + | - | + | Positive | **Positive** | Concordant |
| KP100 | N/A | N/A | 31.57 | **Negative** | + | - | - | Negative | | | | | **Negative** | Concordant |
| KP101 | **25.13** | **25.09** | **29.78** | **Positive** | + | + | + | Positive | | | | | **Positive** | Concordant |
| KP102 | **17.3** | **17.32** | **23.31** | **Positive** | + | + | + | Positive | | | | | **Positive** | Concordant |
| KP103 | **20.52** | **21.35** | **30.83** | **Positive** | + | + | + | Positive | | | | | **Positive** | Concordant |
| KP104 | N/A | N/A | 29.53 | **Negative** | + | - | - | Negative | | | | | **Negative** | Concordant |
| KP105 | 37.23 | N/A | 31.35 | **Negative** | + | - | - | Negative | | | | | **Negative** | Concordant |
| KP106 | 37.95 | N/A | 32.72 | **Negative** | + | - | - | Negative | | | | | **Negative** | Concordant |
| KP107 | **22.99** | **23.49** | **28.44** | **Positive** | + | + | + | Positive | | | | | **Positive** | Concordant |
| KP108 | **29.26** | **30.06** | **28.27** | **Positive** | + | + | + | Positive | | | | | **Positive** | Concordant |
| KP109 | **21.14** | **20.53** | **23.07** | **Positive** | + | + | + | Positive | | | | | **Positive** | Concordant |
| KP110 | N/A | N/A | 20.7 | **Negative** | + | + | - | REPEAT | + | - | - | Negative | **Negative** | Concordant |
| KP111 | N/A | N/A | 20.74 | **Negative** | + | - | - | Negative | | | | | **Negative** | Concordant |
| KP112 | N/A | N/A | 18.7 | **Negative** | + | - | - | Negative | | | | | **Negative** | Concordant |
| KP113 | **24.04** | **23.55** | **20.84** | **Positive** | + | + | + | Positive | | | | | **Positive** | Concordant |
| KP114 | **13.46** | **13.33** | **21.92** | **Positive** | + | + | + | Positive | | | | | **Positive** | Concordant |
| KP115 | 37.97 | 39.68 | 21.46 | **Negative** | + | + | - | REPEAT | + | - | - | Negative | **Negative** | Concordant |
| KP116 | 36.1 | N/A | 23.21 | **Negative** | + | - | + | REPEAT | + | - | - | Negative | **Negative** | Concordant |
| KP117 | **29.51** | **29.41** | **18.39** | **Positive** | + | + | + | Positive | | | | | **Positive** | Concordant |
| KP118 | **15.97** | **15.76** | **24.56** | **Positive** | + | + | + | Positive | | | | | **Positive** | Concordant |
| KP119 | 36.46 | N/A | 24.14 | **Negative** | + | - | - | Negative | | | | | **Negative** | Concordant |
| KP120 | 37.51 | N/A | 23.13 | **Negative** | + | - | - | Negative | | | | | **Negative** | Concordant |
| KP121 | **16.17** | **16.09** | **24.51** | **Positive** | + | + | + | Positive | | | | | **Positive** | Concordant |
| KP122 | **15.78** | **15.67** | **24.33** | **Positive** | + | + | + | Positive | | | | | **Positive** | Concordant |
| KP123 | **21.91** | **21.35** | **31.3** | **Positive** | + | + | + | Positive | | | | | **Positive** | Concordant |
| KP124 | **22.75** | **22.06** | **31.72** | **Positive** | + | + | + | Positive | | | | | **Positive** | Concordant |
| KP125 | **21.41** | **20.8** | **31.43** | **Positive** | + | + | + | Positive | | | | | **Positive** | Concordant |
| KP126 | **18.16** | **17.82** | **23.18** | **Positive** | + | + | + | Positive | | | | | **Positive** | Concordant |
| KP127 | **18.38** | **17.95** | **23.24** | **Positive** | + | + | + | Positive | | | | | **Positive** | Concordant |

| | | | | | | | | | | | | | | |
|---|---|---|---|---|---|---|---|---|---|---|---|---|---|---|
| KP128 | **18.11** | **18.05** | **25.74** | **Positive** | + | + | + | Positive | | | | | **Positive** | Concordant |
| KP129 | **17.75** | **17.92** | **25.86** | **Positive** | + | + | + | Positive | | | | | **Positive** | Concordant |
| KP130 | **20.31** | **20** | **26.32** | **Positive** | + | + | + | Positive | | | | | **Positive** | Concordant |
| KP131 | **22.19** | **21.78** | **23.65** | **Positive** | + | + | + | Positive | | | | | **Positive** | Concordant |
| KP132 | **19.68** | **19.63** | **28.33** | **Positive** | + | + | + | Positive | | | | | **Positive** | Concordant |
| KP133 | **22.71** | **22.63** | **26.05** | **Positive** | + | + | + | Positive | | | | | **Positive** | Concordant |
| KP134 | **22.92** | **22.83** | **25.76** | **Positive** | + | + | + | Positive | | | | | **Positive** | Concordant |
| KP135 | **23.29** | **23.34** | **26.3** | **Positive** | + | + | + | Positive | | | | | **Positive** | Concordant |
| KP136 | 37.98 | 37.52 | 22.14 | **Negative** | + | - | - | Negative | | | | | **Negative** | Concordant |
| KP137 | 37.73 | N/A | 21.61 | **Negative** | + | - | - | Negative | | | | | **Negative** | Concordant |
| KP138 | 36.57 | N/A | 23.71 | **Negative** | + | - | - | Negative | | | | | **Negative** | Concordant |
| KP139 | N/A | N/A | 25.02 | **Negative** | + | - | - | Negative | | | | | **Negative** | Concordant |
| KP140 | N/A | N/A | 27.3 | **Negative** | + | - | - | Negative | | | | | **Negative** | Concordant |
| KP141 | 36.45 | N/A | 25.87 | **Negative** | + | - | - | Negative | | | | | **Negative** | Concordant |
| KP142 | 36.87 | N/A | 25.14 | **Negative** | + | - | - | Negative | | | | | **Negative** | Concordant |
| KP143 | 36.52 | 37.3 | 27.15 | **Negative** | + | - | - | Negative | | | | | **Negative** | Concordant |
| KP144 | 38.58 | N/A | 26.59 | **Negative** | + | - | - | Negative | | | | | **Negative** | Concordant |
| KP145 | 37.6 | 39.87 | 28.71 | **Negative** | + | - | - | Negative | | | | | **Negative** | Concordant |
| KP146 | N/A | N/A | 27.35 | **Negative** | + | - | - | Negative | | | | | **Negative** | Concordant |
| KP147 | 35.47 | | 22.3 | **Negative** | + | - | - | Negative | | | | | **Negative** | Concordant |
| KP148 | 35.08 | 38.31 | 20.77 | **Negative** | + | - | - | Negative | | | | | **Negative** | Concordant |
| KP149 | N/A | N/A | 28.56 | **Negative** | + | - | - | Negative | | | | | **Negative** | Concordant |
| KP150 | 35.84 | 36.55 | 35.29 | **Negative** | + | - | - | Negative | | | | | **Negative** | Concordant |
| KP151 | 31.35 | 31.24 | 26.78 | **Positive** | + | + | + | Positive | | | | | **Positive** | Concordant |
| KP152 | 29.38 | 29.32 | 32.2 | **Positive** | + | - | + | Repeat | + | - | + | Positive | **Positive** | Concordant |
| KP153 | 32.24 | 32.03 | 29.92 | **Positive** | + | + | + | Positive | | | | | **Positive** | Concordant |
| KP154 | 32.28 | 32.24 | 29.29 | **Positive** | + | + | + | Positive | | | | | **Positive** | Concordant |
| KP155 | 24.14 | 24.48 | 27.7 | **Positive** | + | + | + | Positive | | | | | **Positive** | Concordant |
| KP156 | NA | NA | 28.17 | **Negative** | + | - | - | Negative | | | | | **Negative** | Concordant |
| KP157 | NA | NA | 27.4 | **Negative** | + | - | - | Negative | | | | | **Negative** | Concordant |
| KP158 | 24.18 | 24.35 | 27.57 | **Positive** | + | + | + | Positive | | | | | **Positive** | Concordant |
| KP159 | 24.15 | 24.28 | 27.55 | **Positive** | + | + | + | Positive | | | | | **Positive** | Concordant |
| KP160 | 33.04 | 32.63 | 27.71 | **Positive** | + | + | + | Positive | | | | | **Positive** | Concordant |

| | | | | | | | | | | | | | | |
|---|---|---|---|---|---|---|---|---|---|---|---|---|---|---|
| KP161 | 33.6 | 33.74 | 27.3 | Positive | + | - | + | Repeat | + | - | + | Positive | Positive | Concordant |
| KP162 | 31.58 | 31.56 | 26.32 | Positive | + | - | + | Repeat | + | - | + | Positive | Positive | Concordant |
| KP163 | NA | NA | 30.22 | Negative | + | - | - | Negative | | | | | Negative | Concordant |
| KP164 | 31.45 | 31.28 | 25.61 | Positive | + | - | + | Repeat | + | - | + | Positive | Positive | Concordant |
| KP165 | 29.41 | 31.36 | 29.15 | Positive | + | + | + | Positive | | | | | Positive | Concordant |
| KP166 | 27.05 | 27.87 | 25.48 | Positive | + | + | + | Positive | | | | | Positive | Concordant |
| KP167 | 29.26 | 30.06 | 28.27 | Positive | + | + | + | Positive | | | | | Positive | Concordant |
| KP168 | 24.04 | 23.55 | 20.84 | Positive | + | + | + | Positive | | | | | Positive | Concordant |
| KP169 | 29.51 | 29.41 | 18.39 | Positive | + | - | + | Repeat | + | - | + | Positive | Positive | Concordant |
| KP170 | 21.14 | 20.53 | 23.07 | Positive | + | + | + | Positive | | | | | Positive | Concordant |
| KP171 | NA | NA | 32.21 | Negative | + | - | - | Negative | | | | | Negative | Concordant |
| KP172 | NA | NA | 30.49 | Negative | + | - | - | Negative | | | | | Negative | Concordant |
| KP173 | 24.13 | 24.14 | 29.21 | Positive | + | - | + | Repeat | + | - | + | Positive | Positive | Concordant |
| KP174 | 22.78 | 25.76 | 26.04 | Positive | - | + | + | Positive | | | | | Positive | Concordant |
| KP175 | 22.69 | 26.97 | 22.8 | Positive | + | - | + | Repeat | + | - | + | Positive | Positive | Concordant |
| KP176 | 17.3 | 17.32 | 23.31 | Positive | + | + | + | Positive | | | | | Positive | Concordant |
| KP177 | 20.52 | 21.35 | 30.83 | Positive | + | + | + | Positive | | | | | Positive | Concordant |
| KP178 | 13.46 | 13.33 | 21.92 | Positive | + | + | + | Positive | | | | | Positive | Concordant |
| KP179 | 26.25 | 27.5 | 25.47 | Positive | + | + | + | Positive | | | | | Positive | Concordant |
| KP180 | 15.97 | 15.76 | 24.56 | Positive | + | + | + | Positive | | | | | Positive | Concordant |
| KP181 | NA | NA | 28.94 | Negative | + | - | - | Negative | | | | | Negative | Concordant |
| KP182 | NA | NA | 30.27 | Negative | + | - | - | Negative | | | | | Negative | Concordant |
| KP183 | NA | NA | 31 | Negative | + | - | - | Negative | | | | | Negative | Concordant |
| KP184 | 16.17 | 16.09 | 24.51 | Positive | + | + | + | Positive | | | | | Positive | Concordant |
| KP185 | 21.91 | 21.35 | 31.3 | Positive | + | + | + | Positive | | | | | Positive | Concordant |
| KP186 | 22.75 | 22.06 | 31.72 | Positive | + | + | + | Positive | | | | | Positive | Concordant |
| KP187 | 21.41 | 20.88 | 31.43 | Positive | + | + | + | Positive | | | | | Positive | Concordant |
| KP188 | 18.16 | 17.82 | 23.18 | Positive | + | + | + | Positive | | | | | Positive | Concordant |
| KP189 | 18.38 | 17.95 | 23.24 | Positive | + | + | + | Positive | | | | | Positive | Concordant |
| KP190 | 18.11 | 18.05 | 25.74 | Positive | + | + | + | Positive | | | | | Positive | Concordant |
| KP191 | NA | NA | 30.49 | Negative | + | - | - | Negative | | | | | Negative | Concordant |
| KP192 | NA | NA | 28.76 | Negative | + | - | - | Negative | | | | | Negative | Concordant |
| KP193 | 17.75 | 17.92 | 25.86 | Positive | + | + | + | Positive | | | | | Positive | Concordant |

| | | | | | | | | | | | | | | |
|---|---|---|---|---|---|---|---|---|---|---|---|---|---|---|
| KP194 | 20.21 | 20 | 26.32 | **Positive** | + | + | + | Positive | | | | | **Positive** | Concordant |
| KP195 | 22.19 | 21.78 | 23.65 | **Positive** | + | + | + | Positive | | | | | **Positive** | Concordant |
| KP196 | 19.68 | 19.63 | 28.33 | **Positive** | + | + | + | Positive | | | | | **Positive** | Concordant |
| KP197 | 24.41 | 24.99 | 18.57 | **Positive** | + | + | + | Positive | | | | | **Positive** | Concordant |
| KP198 | 22.71 | 22.63 | 26.05 | **Positive** | + | + | + | Positive | | | | | **Positive** | Concordant |
| KP199 | 22.92 | 22.83 | 25.76 | **Positive** | + | + | + | Positive | | | | | **Positive** | Concordant |
| KP200 | 23.29 | 23.34 | 26.3 | **Positive** | + | + | + | Positive | | | | | **Positive** | Concordant |

**Supplementary Table S2**: Accession number of complete genome sequences of SARS CoV-2 used for sequence alignment and bioinformatics

| | | | | | |
|---|---|---|---|---|---|
| NC_045512.2 | MW054108.1 | MT873103.1 | MT609564.1 | MT951955.1 | MW084412.1 |
| MT679209.1 | MW052624.1 | MT607246.1 | MT873390.1 | MT627419.1 | MW084547.1 |
| MW181540.1 | MT873369.1 | MT657275.1 | MT873211.1 | MT704119.1 | MT951928.1 |
| MT679197.1 | MT873174.1 | LC556316.1 | MT873185.1 | MT873200.1 | MW172728.1 |
| MW181431.1 | MT873286.1 | MT656256.1 | MT873451.1 | MT731746.1 | MW084506.1 |
| MT873406.1 | MT873319.1 | MT664105.1 | MT873160.1 | MW053874.1 | MW172559.1 |
| MT635644.1 | MW054103.1 | MT656010.1 | MT873334.1 | MT843312.1 | MW084576.1 |
| MT873171.1 | MW052620.1 | MT656590.1 | MT873115.1 | MW084559.1 | MT782360.1 |
| MT873483.1 | MT827246.1 | MT627415.1 | MT873488.1 | MT739433.1 | MT877387.1 |
| MT577628.1 | MW346348.1 | MT890462.1 | MT873290.1 | MT913046.1 | MT913094.1 |
| MT873374.1 | MW053935.1 | LC556320.1 | MT873119.1 | MT755593.1 | MW172561.1 |
| MT873912.1 | MW047312.1 | MT873257.1 | MT873473.1 | MT843288.1 | MW084507.1 |
| MT873316.1 | MT951962.1 | MT956642.1 | MT873111.1 | MT873201.1 | MW053752.1 |
| MT951954.1 | MW346179.1 | MT627426.1 | MT873093.1 | MT782371.1 | MW241190.1 |
| MT806882.1 | MT843320.1 | MT872498.1 | MT731292.1 | MW053869.1 | MT755596.1 |
| MT806881.1 | MW342708.1 | MT750460.1 | MT873367.1 | MT873803.1 | MW054107.1 |
| MT873181.1 | MT750335.1 | MT704127.1 | MT873360.1 | MT679163.1 | MW052623.1 |
| MT782348.1 | MT609575.1 | MT664107.1 | MT873177.1 | MT679170.1 | MW053804.1 |
| MW084536.1 | MT609571.1 | MT873275.1 | MT873487.1 | MW290942.1 | MW054105.1 |
| MT782361.1 | MT750440.1 | MW181518.1 | MT873362.1 | MT679161.1 | MW052622.1 |

| | | | | | |
|---|---|---|---|---|---|
| MW346345.1 | MT750439.1 | MT671821.1 | MT873284.1 | MT913113.1 | MW054106.1 |
| MT782364.1 | MT750446.1 | MT627427.1 | MT873294.1 | MT739431.1 | MW084535.1 |
| MT913137.1 | MT750436.1 | MT679192.1 | MT873355.1 | MT913044.1 | MW084549.1 |
| MT913110.1 | MT750366.1 | MT873062.1 | MT873225.1 | MT913133.1 | MT877372.1 |
| MT782381.1 | MT750367.1 | MT873245.1 | MT873393.1 | MT913101.1 | MT782372.1 |
| MT635649.1 | MW210975.1 | MT755889.1 | MT873318.1 | MW084441.1 | MT806874.1 |
| MT782377.1 | MT750393.1 | MT755883.1 | MT873207.1 | MT755599.1 | MW084574.1 |
| MT764170.1 | MT750402.1 | MT755899.1 | MT873383.1 | MW053883.1 | MT782349.1 |
| MT873240.1 | MT750412.1 | MT755885.1 | MT873258.1 | MT873469.1 | MT782355.1 |
| MT873446.1 | MT750339.1 | MT755887.1 | MT873157.1 | MT992739.1 | MT877427.1 |
| MT873391.1 | MW031799.1 | MT755884.1 | MT873314.1 | MW053925.1 | MT877414.1 |
| MT873192.1 | MW031800.1 | MT755886.1 | MT873233.1 | MW210993.1 | MT873917.1 |
| MT873346.1 | MW031801.1 | MT755898.1 | MT873097.1 | MT739426.1 | MT755597.1 |
| MT873285.1 | MW031802.1 | MT755890.1 | MT873401.1 | MT913039.1 | MT782362.1 |
| MT873053.1 | MT873152.1 | MT755896.1 | MT873442.1 | MT913105.1 | MW053973.1 |
| MT873203.1 | MT873050.1 | MT755894.1 | MT873088.1 | MT755602.1 | MW346350.1 |
| MT873327.1 | MT750433.1 | MT755891.1 | MT873105.1 | MT653099.1 | MW053945.1 |
| MT873221.1 | MT750444.1 | MT755892.1 | MT873165.1 | MW053878.1 | MW084550.1 |
| MT873470.1 | MT738101.1 | MT755897.1 | MT873414.1 | MW134008.1 | MT873806.1 |
| MT873067.1 | MT655742.1 | MT755893.1 | MT873247.1 | MW084558.1 | MW053946.1 |
| MT873135.1 | MT657271.1 | MT755888.1 | MT873179.1 | MW053885.1 | MW047311.1 |
| MT873159.1 | MT775833.1 | MT755895.1 | MT873123.1 | MW084440.1 | MW047307.1 |
| MT873404.1 | MT775830.1 | MT873259.1 | MT873496.1 | MT873864.1 | MW047316.1 |
| MT873188.1 | MW255832.1 | MT873324.1 | MT873323.1 | MT873120.1 | MT627432.1 |
| MT873453.1 | MT671828.1 | MT929083.1 | MT873418.1 | MT873169.1 | MT955360.1 |
| MT873076.1 | MW031803.1 | MT609565.1 | MT764166.1 | MT873098.1 | MT956918.1 |
| MT951941.1 | MT873405.1 | MT873120.1 | MT627432.1 | MT627429.1 | MW047311.1 |
| MT929095.1 | MT873421.1 | MT873169.1 | MT955360.1 | MT873139.1 | MW047307.1 |
| MT827216.1 | MT873218.1 | MT873098.1 | MT956918.1 | MT873108.1 | MW047316.1 |
| MW210997.1 | MT873086.1 | MT873268.1 | MT577009.1 | MW053929.1 | MT951955.1 |
| MT873457.1 | MT873128.1 | MT873063.1 | MT577010.1 | MT873353.1 | MT627419.1 |

| | | | | | |
|---|---|---|---|---|---|
| MW047315.1 | MT873104.1 | MT951965.1 | MT956912.1 | MW241191.1 | MT704119.1 |
| MT873134.1 | MT873438.1 | MW241201.1 | MT956917.1 | MT873396.1 | MT873200.1 |
| MT806884.1 | MT873287.1 | MT739466.1 | MW010236.1 | MT679159.1 | MT731746.1 |
| MT609566.1 | MT873217.1 | MT913079.1 | MW210979.1 | MT704123.1 | MW053874.1 |
| MT609569.1 | MT873308.1 | MT873447.1 | MW276456.1 | MT873370.1 | MT843312.1 |
| MT609568.1 | MT873464.1 | MT873419.1 | MW276454.1 | MT750455.1 | MW084559.1 |
| MT820131.1 | MT873454.1 | MT873329.1 | MW276451.1 | MW053708.1 | MT739433.1 |
| MT750418.1 | MT873326.1 | MT873506.1 | MW276453.1 | MT873350.1 | MT913046.1 |
| MT750386.1 | MT873254.1 | MT873065.1 | MW054109.1 | MT873153.1 | MT755593.1 |
| MT750423.1 | MT873477.1 | MT873212.1 | MW052625.1 | MT951946.1 | MT843288.1 |
| MT750385.1 | MT873141.1 | MT873452.1 | MW053938.1 | MT873410.1 | MT873201.1 |
| MT679164.1 | MT609581.1 | MT873356.1 | MT913124.1 | MT873458.1 | MT782371.1 |
| MT750421.1 | MT609583.1 | MT873389.1 | MT873167.1 | MT873149.1 | MW053869.1 |
| MT750353.1 | MT609582.1 | MT873321.1 | MT873416.1 | MT873183.1 | MT873803.1 |
| MT750416.1 | LC581365.1 | MT873121.1 | MT820128.1 | MT873333.1 | MT679163.1 |
| MT750352.1 | MT873402.1 | MT873163.1 | MT820127.1 | MT873422.1 | MT679170.1 |
| MT750477.1 | MT873403.1 | MT873095.1 | MT609562.1 | MT873467.1 | MW290942.1 |
| MT750474.1 | MT873272.1 | MT873235.1 | MT843305.1 | MT873424.1 | MT679161.1 |
| MT750478.1 | MW011764.1 | MT873311.1 | MT873052.1 | MT873069.1 | MT913113.1 |
| MT750465.1 | MW011768.1 | MT873347.1 | MT873060.1 | MT873100.1 | MT739431.1 |
| MT750470.1 | MT671817.1 | MT873263.1 | MT951974.1 | MT628262.1 | MT913044.1 |
| MT750469.1 | MW011766.1 | MT680235.1 | MT609588.1 | MW047308.1 | MT913133.1 |
| MT750482.1 | MW011767.1 | MT873505.1 | MT913123.1 | MT671827.1 | MT913101.1 |
| MT873208.1 | MW011763.1 | MT873195.1 | MT913125.1 | MT627428.1 | MW084441.1 |
| MT873348.1 | MW011765.1 | MT873204.1 | MT609559.1 | MT873073.1 | MT755599.1 |
| MT956913.1 | MW011762.1 | MT873190.1 | MT820132.1 | MT873099.1 | MW053883.1 |
| MT956916.1 | MT873156.1 | MT873501.1 | MT820130.1 | MT731764.1 | MT873469.1 |
| MT956911.1 | MT873296.1 | MT873236.1 | MT873465.1 | MT733120.1 | MT992739.1 |
| MT956915.1 | MT873261.1 | MT873230.1 | MT609600.1 | MT627420.1 | MW053925.1 |
| MT873143.1 | MT873147.1 | MT873303.1 | MT873140.1 | MT627411.1 | MW210993.1 |
| MT873397.1 | MT873377.1 | MT873220.1 | MT627435.1 | MT627424.1 | MT739426.1 |

| | | | | | |
|---|---|---|---|---|---|
| MT827233.1 | MT873409.1 | MT873430.1 | MT750501.1 | MT731285.1 | MT913039.1 |
| MT873262.1 | MT873429.1 | MW346370.1 | MT873249.1 | MT679205.1 | MT913105.1 |
| MT731327.1 | MT873462.1 | MT873399.1 | MT873082.1 | MT627431.1 | MT755602.1 |
| MT731673.1 | MT873243.1 | MT873338.1 | MT873256.1 | MT679206.1 | MT653099.1 |
| MT873124.1 | MT873051.1 | MT873295.1 | MT873279.1 | MT873292.1 | MW053878.1 |
| MT873084.1 | MT873336.1 | MT873448.1 | MW047309.1 | MT680229.1 | MW134008.1 |
| MT873158.1 | MW047317.1 | MT873094.1 | MW047318.1 | MT911805.1 | MW084558.1 |
| MT873299.1 | MT873116.1 | MT873222.1 | MT609560.1 | MW241204.1 | MW053885.1 |
| MT873330.1 | MT873131.1 | MW047314.1 | MT820129.1 | MW004168.1 | MW084440.1 |
| MT873309.1 | MT873058.1 | MW047310.1 | MW047313.1 | MT731468.1 | MT873864.1 |
| MW084550.1 | MT679197.1 | MT873174.1 | LC556316.1 | MT873185.1 | MW210997.1 |
| MT873806.1 | MW181431.1 | MT873286.1 | MT656256.1 | MT873451.1 | MT873457.1 |
| MW053946.1 | MT873406.1 | MT873319.1 | MT664105.1 | MT873160.1 | MW047315.1 |
| MW084412.1 | MT635644.1 | MW054103.1 | MT656010.1 | MT873334.1 | MT873134.1 |
| MW084547.1 | MT873171.1 | MW052620.1 | MT656590.1 | MT873115.1 | MT806884.1 |
| MT951928.1 | MT873483.1 | MT827246.1 | MT627415.1 | MT873488.1 | MT609566.1 |
| MW172728.1 | MT577628.1 | MW346348.1 | MT890462.1 | MT873290.1 | MT609569.1 |
| MW084506.1 | MT873374.1 | MW053935.1 | LC556320.1 | MT873119.1 | MT609568.1 |
| MW172559.1 | MT873912.1 | MW047312.1 | MT873257.1 | MT873473.1 | MT820131.1 |
| MW084576.1 | MT873316.1 | MT951962.1 | MT956642.1 | MT873111.1 | MT750418.1 |
| MT782360.1 | MT951954.1 | MW346179.1 | MT627426.1 | MT873093.1 | MT750386.1 |
| MT877387.1 | MT806882.1 | MT843320.1 | MT872498.1 | MT731292.1 | MT750423.1 |
| MT913094.1 | MT806881.1 | MW342708.1 | MT750460.1 | MT873367.1 | MT750385.1 |
| MW172561.1 | MT873181.1 | MT750335.1 | MT704127.1 | MT873360.1 | MT679164.1 |
| MW084507.1 | MT782348.1 | MT609575.1 | MT664107.1 | MT873177.1 | MT750421.1 |
| MW053752.1 | MW084536.1 | MT609571.1 | MT873275.1 | MT873487.1 | MT750353.1 |
| MW241190.1 | MT782361.1 | MT750440.1 | MW181518.1 | MT873362.1 | MT750416.1 |
| MT755596.1 | MW346345.1 | MT750439.1 | MT671821.1 | MT873284.1 | MT750352.1 |
| MW054107.1 | MT782364.1 | MT750446.1 | MT627427.1 | MT873294.1 | MT750477.1 |
| MW052623.1 | MT913137.1 | MT750436.1 | MT679192.1 | MT873355.1 | MT750474.1 |
| MW053804.1 | MT913110.1 | MT750366.1 | MT873062.1 | MT873225.1 | MT750478.1 |

| | | | | | |
|---|---|---|---|---|---|
| MW054105.1 | MT782381.1 | MT750367.1 | MT873245.1 | MT873393.1 | MT750465.1 |
| MW052622.1 | MT635649.1 | MW210975.1 | MT755889.1 | MT873318.1 | MT750470.1 |
| MW054106.1 | MT782377.1 | MT750393.1 | MT755883.1 | MT873207.1 | MT750469.1 |
| MW084535.1 | MT764170.1 | MT750402.1 | MT755899.1 | MT873383.1 | MT750482.1 |
| MW084549.1 | MT873240.1 | MT750412.1 | MT755885.1 | MT873258.1 | MT873208.1 |
| MT877372.1 | MT873446.1 | MT750339.1 | MT755887.1 | MT873157.1 | MT873348.1 |
| MT782372.1 | MT873391.1 | MW031799.1 | MT755884.1 | MT873314.1 | MT956913.1 |
| MT806874.1 | MT873192.1 | MW031800.1 | MT755886.1 | MT873233.1 | MT956916.1 |
| MW084574.1 | MT873346.1 | MW031801.1 | MT755898.1 | MT873097.1 | MT956911.1 |
| MT782349.1 | MT873285.1 | MW031802.1 | MT755890.1 | MT873401.1 | MT956915.1 |
| MT782355.1 | MT873053.1 | MT873152.1 | MT755896.1 | MT873442.1 | MT873143.1 |
| MT877427.1 | MT873203.1 | MT873050.1 | MT755894.1 | MT873088.1 | MT873397.1 |
| MT877414.1 | MT873327.1 | MT750433.1 | MT755891.1 | MT873105.1 | MT827233.1 |
| MT873917.1 | MT873221.1 | MT750444.1 | MT755892.1 | MT873165.1 | MT873262.1 |
| MT755597.1 | MT873470.1 | MT738101.1 | MT755897.1 | MT873414.1 | MT731327.1 |
| MT782362.1 | MT873067.1 | MT655742.1 | MT755893.1 | MT873247.1 | MT731673.1 |
| MW053973.1 | MT873135.1 | MT657271.1 | MT755888.1 | MT873179.1 | MT873124.1 |
| MW346350.1 | MT873159.1 | MT775833.1 | MT755895.1 | MT873123.1 | MT873084.1 |
| MW053945.1 | MT873404.1 | MT775830.1 | MT873259.1 | MT873496.1 | MT873158.1 |
| NC_045512.2 | MT873188.1 | MW255832.1 | MT873324.1 | MT873323.1 | MT873299.1 |
| MT679209.1 | MT873453.1 | MT671828.1 | MT929083.1 | MT873418.1 | MT873330.1 |
| MW181540.1 | MT873076.1 | MW031803.1 | MT609565.1 | MT764166.1 | MT873309.1 |
| MT627429.1 | MW054108.1 | MT873103.1 | MT609564.1 | MT951941.1 | MT873405.1 |
| MT873139.1 | MW052624.1 | MT607246.1 | MT873390.1 | MT929095.1 | MT873421.1 |
| MT873108.1 | MT873369.1 | MT657275.1 | MT873211.1 | MT827216.1 | MT873218.1 |
| MT873086.1 | MT873268.1 | MT577009.1 | MW053929.1 | | |
| MT873128.1 | MT873063.1 | MT577010.1 | MT873353.1 | | |
| MT873104.1 | MT951965.1 | MT956912.1 | MW241191.1 | | |
| MT873438.1 | MW241201.1 | MT956917.1 | MT873396.1 | | |
| MT873287.1 | MT739466.1 | MW010236.1 | MT679159.1 | | |
| MT873217.1 | MT913079.1 | MW210979.1 | MT704123.1 | | |

| | | | |
|---|---|---|---|
| MT873308.1 | MT873447.1 | MW276456.1 | MT873370.1 |
| MT873464.1 | MT873419.1 | MW276454.1 | MT750455.1 |
| MT873454.1 | MT873329.1 | MW276451.1 | MW053708.1 |
| MT873326.1 | MT873506.1 | MW276453.1 | MT873350.1 |
| MT873254.1 | MT873065.1 | MW054109.1 | MT873153.1 |
| MT873477.1 | MT873212.1 | MW052625.1 | MT951946.1 |
| MT873141.1 | MT873452.1 | MW053938.1 | MT873410.1 |
| MT609581.1 | MT873356.1 | MT913124.1 | MT873458.1 |
| MT609583.1 | MT873389.1 | MT873167.1 | MT873149.1 |
| MT609582.1 | MT873321.1 | MT873416.1 | MT873183.1 |
| LC581365.1 | MT873121.1 | MT820128.1 | MT873333.1 |
| MT873402.1 | MT873163.1 | MT820127.1 | MT873422.1 |
| MT873403.1 | MT873095.1 | MT609562.1 | MT873467.1 |
| MT873272.1 | MT873235.1 | MT843305.1 | MT873424.1 |
| MW011764.1 | MT873311.1 | MT873052.1 | MT873069.1 |
| MW011768.1 | MT873347.1 | MT873060.1 | MT873100.1 |
| MT671817.1 | MT873263.1 | MT951974.1 | MT628262.1 |
| MW011766.1 | MT680235.1 | MT609588.1 | MW047308.1 |
| MW011767.1 | MT873505.1 | MT913123.1 | MT671827.1 |
| MW011763.1 | MT873195.1 | MT913125.1 | MT627428.1 |
| MW011765.1 | MT873204.1 | MT609559.1 | MT873073.1 |
| MW011762.1 | MT873190.1 | MT820132.1 | MT873099.1 |
| MT873156.1 | MT873501.1 | MT820130.1 | MT731764.1 |
| MT873296.1 | MT873236.1 | MT873465.1 | MT733120.1 |
| MT873261.1 | MT873230.1 | MT609600.1 | MT627420.1 |
| MT873147.1 | MT873303.1 | MT873140.1 | MT627411.1 |
| MT873377.1 | MT873220.1 | MT627435.1 | MT627424.1 |
| MT873409.1 | MT873430.1 | MT750501.1 | MT731285.1 |
| MT873429.1 | MW346370.1 | MT873249.1 | MT679205.1 |
| MT873462.1 | MT873399.1 | MT873082.1 | MT627431.1 |
| MT873243.1 | MT873338.1 | MT873256.1 | MT679206.1 |

| | | | |
|---|---|---|---|
| MT873051.1 | MT873295.1 | MT873279.1 | MT873292.1 |
| MT873336.1 | MT873448.1 | MW047309.1 | MT680229.1 |
| MW047317.1 | MT873094.1 | MW047318.1 | MT911805.1 |
| MT873116.1 | MT873222.1 | MT609560.1 | MW241204.1 |
| MT873131.1 | MW047314.1 | MT820129.1 | MW004168.1 |
| MT873058.1 | MW047310.1 | MW047313.1 | MT731468.1 |
